# Internal tremors and vibrations in long COVID: a cross-sectional study

**DOI:** 10.1101/2023.06.19.23291598

**Authors:** Tianna Zhou, Mitsuaki Sawano, Adith S. Arun, César Caraballo, Teresa Michelsen, Lindsay McAlpine, Bornali Bhattacharjee, Yuan Lu, Rohan Khera, Chenxi Huang, Frederick Warner, Akiko Iwasaki, Harlan M. Krumholz

## Abstract

**Importance:** Internal tremors and vibrations symptoms have been described as part of neurologic disorders but not fully described as a part of long COVID.

**Objective:** To compare demographics, socioeconomic characteristics, pre-pandemic comorbidities, new-onset conditions, and long COVID symptoms between people with internal tremors and vibrations as part of their long COVID symptoms and people with long COVID but without these symptoms.

**Design:** A cross-sectional study, Listen to Immune, Symptom and Treatment Experiences Now (LISTEN), of adults with and without long COVID and post-vaccination syndrome, defined by self-report.

**Setting:** Hugo Health Kindred, a decentralized digital research platform hosting a network of English-speaking adults interested in contributing to COVID-related research. No geographic limitation applied.

**Participants:** The study population included 423 participants who enrolled in LISTEN between May 2022 and June 2023, completed the initial and the conditions and symptoms surveys, reported long COVID, and did not report post-vaccination syndrome.

**Exposure:** Long COVID symptoms of internal tremors and vibrations.

**Main outcomes and Measures:** Demographics, pre-pandemic comorbidities, and current conditions, other symptoms, and quality of life at the time of surveys.

**Results:** Of the 423 participants (median age, 46 years [IQR, 38-56]), 74% were female, 87% were Non-Hispanic White, 92% lived in the United States, 46% were infected before the Delta wave, and 158 (37%) reported “internal tremors, or buzzing/vibration” as a long COVID symptom. Before long COVID, the groups had similar comorbidities. Participants with internal tremors were different from others in having worse health as measured by the Euro-QoL visual analogue scale (median: 40 points [IQR, 30-60] vs. 50 points [IQR, 35-62], P = 0.007), having financial difficulties caused by the pandemic (very much financial difficulties, 22% [95% CI, 16-30] vs. 11% [7.3-15], P < 0.001), often feeling socially isolated (43% [95% CI, 35-52] vs. 37% [31-43], P = 0.039), and having higher rates of self-reported new-onset mast cell disorders (11% [95% CI, 7.1-18] vs. 2.6% [1.2-5.6], Bonferroni-adjusted P = 0.008) and neurologic conditions (including but not limited to seizures, dementia, multiple sclerosis, Parkinson’s disease, neuropathy, etc.; 22% [95% CI, 16-29] vs. 8.3% [5.4-12], Bonferroni-adjusted P = 0.004).

**Conclusions and Relevance:** Among people with long COVID, those with internal tremors and vibrations have several other associated symptoms and worse health status, despite having similar pre-pandemic comorbidities, suggesting it may reflect a severe phenotype of long COVID.

**KEY POINTS:** *Question:* Do people with long COVID symptoms of internal tremors and vibrations differ from others with long COVID but without these symptoms?

*Findings:* In this cross-sectional study that included 423 adults with long COVID, 158 (37%) reported having “internal tremors, or buzzing/vibration,” had worse quality of life, more financial difficulties, and higher rates of new-onset mast cell disorders and neurologic conditions, compared with others with long COVID but without internal tremors and vibrations.

*Meaning:* Internal tremors and vibrations may reflect a severe phenotype of long COVID.

## INTRODUCTION

Internal tremors and vibrations are an understudied symptom, despite their first written descriptions by a patient in the 1800s.^1^ Internal tremors are described as a movement or sensation of movement at any location inside the body. They can occur either with or without visible external movement or muscle spasms.^2, 3^ Until recently, internal tremors were mostly described in Parkinson’s disease and essential tremor.^1–6^ Among patients with Parkinson’s disease and essential tremor, internal tremors have been associated with anxiety and sensory abnormalities such as aching, tingling, and burning.^5^ In a qualitative study, we described long COVID symptoms of internal tremors and vibrations and their substantial and negative impact on people’s quality of life.^2^ Symptoms potentially related to internal tremors, such as tremors, abnormal movements, and numbness/tingling, were described in the Researching COVID to Enhance Recovery (RECOVER) study of a large prospective cohort of people with long COVID; abnormal movements contributed to a preliminary symptom score for defining long COVID.^7^

Herein, we compared the demographics, socioeconomic characteristics, pre-pandemic comorbidities, and new-onset conditions between people with long COVID symptoms of internal tremors and vibrations and others with long COVID. In addition, we built a model to classify the presence and absence of internal tremors based on participants’ other long COVID symptoms. Throughout this paper, terminology and definitions were deliberately chosen to respect participants’ epistemic authority. For example, long COVID was defined by participants’ self-report rather than definitions proposed by public health and medical organizations.^8^ Similarly, this paper used the term “internal tremors,” a shortened phrase to denote “internal tremors and vibrations,” which reflects the language used by participants to describe their symptom experience. Our previous qualitative study found that some people with internal tremors and vibrations experienced those symptoms in varied anatomic locations and a subset also experienced visible tremors.^2^ Thus, internal tremors were considered to be different from the more widely known definition of tremors as “involuntary, rhythmic, oscillatory movement of a body part,”^9^ which was referred to as “tremors or shakiness” in this study. In this paper’s context, internal tremors were considered a participant-reported symptom rather than a clinician-observed sign.^10^

## METHODS

### Study design

We used data from the long COVID component of the Listen to Immune, Symptom, and Treatment Experiences Now (LISTEN) Study, an online, decentralized, participant-centric, observational study of adults interested in contributing to COVID-related research. The LISTEN Study included two parts: survey and electronic health record data collection and analysis to characterize clinical phenotypes and biospecimen collection for immunophenotyping. This study focused on survey data.

### Study sample

LISTEN recruited from people who joined Hugo Health Kindred, an online network of individuals 18 years and older who were interested in contributing to COVID-related research. The objective of the LISTEN study was to include people with self-reported long COVID, post-vaccination syndrome, and those who could act as controls; this paper focused on people with long COVID. People joined Hugo Health Kindred by word of mouth, including via social media. Additionally, a subset of active Kindred members was invited to join a Kindred Advisory Task Force to help recruit others by sharing information about Kindred. Eligibility criteria to join LISTEN were 1) age 18 years or older and 2) English speaking.

### Data collection

Those who joined Kindred were offered a series of surveys developed in an iterative process that included input from participants. Surveys were available for completion online on computers or mobile devices. Participants were sent electronic reminders to encourage survey completion. The data became part of each person’s data repository, available to be shared with research studies. People in Kindred were offered the opportunity to join LISTEN; e-consent was obtained online. This study was restricted to people who joined LISTEN and had completed their demographic and conditions and symptoms surveys.

Demographic and socioeconomic survey items included age, gender, race and ethnicity, marital status, pre-pandemic employment and income, housing insecurity, and country of residence. Self-reported time of index SARS-CoV-2 infection was categorized as Pre-Delta (before 26 June 2021), Delta (26 June 2021–24 December 2021), Omicron (25 December 2021– 25 June 2022), and Post-Omicron (after 25 June 2022), consistent with time period definitions associated with dominant variants of SARS-CoV-2.^11^ SARS-CoV-2 infection severity was assessed by self-reported hospitalization history for COVID-related conditions.

The demographic survey included a question about health status, assessed by the Euro-QoL visual analogue scale (EQ-VAS), and a question about symptom severity: “We are trying to get a sense of how bad your long COVID symptoms are when you feel them the most. On the slider below, with 0 being a trivial illness and 100 being unbearable, please let us know what the worst days are like” (eMethods 1).^12^

The conditions and symptoms survey assessed pre-pandemic comorbidities, current conditions, and long COVID symptoms. Pre-pandemic comorbidities were assessed using the question, “Have you ever been told by a doctor before January 2020 that you have any of the following?” followed by a list of 38 diagnostic categories, “other,” and “none of the above” (eMethods 2). Current conditions were assessed using the question, “Currently, have you ever been told by a doctor that you have any of the following?” followed by a list of 39 diagnostic categories, “other,” and “none of the above” (eMethods 3). For each participant, we defined new-onset conditions as those reported as a current condition but not reported as a pre-pandemic comorbidity.

Long COVID symptoms were assessed by the question, “Please select all following health conditions that you have had as a result of long COVID,” followed by a list of 96 specific symptoms, “other,” and “none of the above” (eMethods 4). Phrasings of symptoms were created in collaboration with participants and were reported in this study’s tables as they appeared on the survey. We approximated participants’ composite symptom score based on the RECOVER Consortium’s proposed criteria (eMethods 5). Because LISTEN did not use the same survey instruments to assess symptom severity as RECOVER, two types of composite scores (including and excluding symptoms with severity criteria) were calculated.

### Statistical analysis

We described participant characteristics using percentages for categorical variables, and median and interquartile range (IQR) for continuous variables. We compared participants with and without internal tremors on their demographic and socioeconomic characteristics, conditions, and long COVID symptoms. We used chi-squared tests and Fisher’s exact tests to compare responses for categorical variables and Wilcoxon rank-sum tests and Kruskal-Wallis rank-sum tests for continuous variables. When comparing the three domains of pre-pandemic comorbidities, new-onset conditions, and long COVID symptoms between the two groups, we corrected for multiple testing using the Bonferroni method within each domain and reported adjusted P-values. All tests were two-sided. P < 0.05 was considered statistically significant. By using the Bonferroni method, family-wise error rates were controlled at the level of 0.05. All statistical analyses were done in R version 4.2.3 (2023-03-15).

We used feature importance in gradient boosted tree machine learning models to identify the most important symptoms for differentiating participants experiencing internal tremors compared to those not experiencing internal tremors.^13, 14^ We trained models to predict whether a participant experienced internal tremors or not, using information on presence or absence of other symptoms for each participant, with 5-fold 5-repeat cross-validation. We dropped two sex-specific symptom variables related to menstruation. We selected the hyperparameters with highest area under the curve (AUC) from the internal cross-validation. Then, we computed the importance of each variable in differentiating participants with and without internal tremors using a permutation-based approach.^15^ We sorted the variables based on their importance and, using this fixed sorting, progressively excluded those with least importance from the model by evaluating the change in the AUC. We selected the best model and corresponding number of variables when the AUC first decreased by at least 1.5%. If a drop of this magnitude did not occur, we selected the model with the largest drop in AUC.

To assess the robustness of this process to choice of modeling methods and variable importance metrics, this process was repeated with an XGBoost model^16^ with the gain in accuracy metric used to assess variable importance, as well as an XGBoost model with the Shapley value^17^ used to assess variable importance. We compare each of the three methods’ results for variable importance values using Pearson correlation coefficients.

Code to reproduce machine learning analyses and generate the associated figures can be found: https://github.com/aditharun/tremors-ml

### Ethical considerations

The LISTEN study was approved by the Yale University Institutional Review Board on April 1, 2022. Participants were provided electronic written consent forms. LISTEN conforms to the Declaration of Helsinki and STROBE reporting guidelines.

## RESULTS

From May 2022 to June 2023, 614 people with long COVID consented to and enrolled in LISTEN and completed the demographic survey (Figure 1). Among them, 191 participants (31%) were excluded due to incomplete conditions and symptoms surveys, leaving 423 participants (69%) in the study population (Figure 1).

**Figure 1.**
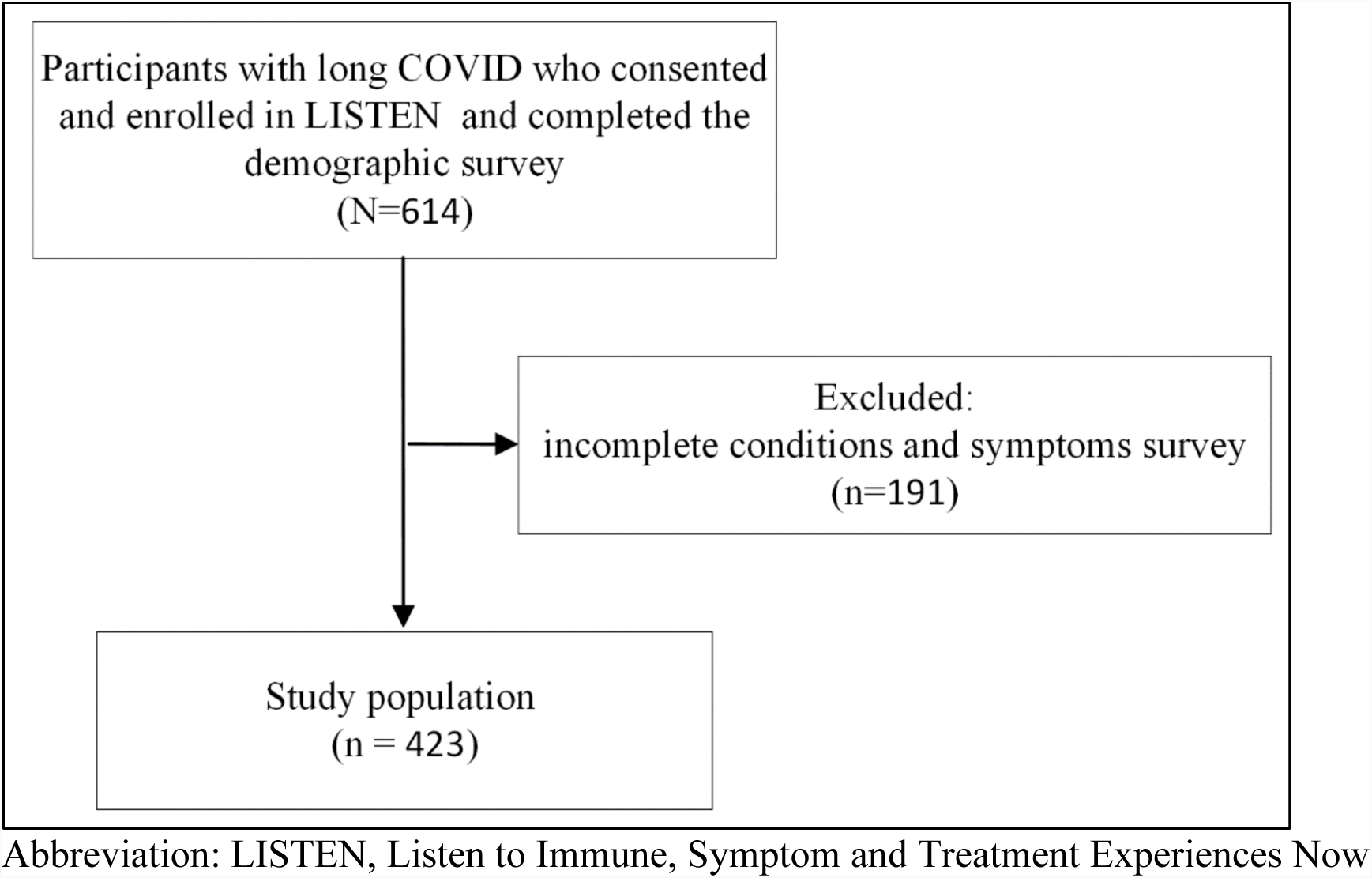
Study population.

### Demographic and pre-pandemic socioeconomic characteristics

Among participants with long COVID, median age was 46 years (IQR, 38-56), 74% were female, 87% were Non-Hispanic White, and 158 participants (37%) reported internal tremors (Table 1). Compared with participants without internal tremors, those with internal tremors were more likely to be female (81% vs. 70%, P = 0.02). The two groups were similar in age, race and ethnicity, marital status, pre-pandemic employment status, and pre-pandemic household income.

**Table 1.** Participant demographics, socioeconomic characteristics, and SARS-CoV-2 infection characteristics.

|  | **Overall, N = 423** | **No internal tremors, N = 265** | **Has internal tremors, N = 158** | **p-value<sup>1</sup>** |
| --- | --- | --- | --- | --- |
|  | **n/N, % [95% CI]** | **n/N, % [95% CI]** | **n/N, % [95% CI]** |  |
| Age (years), median, IQR | 46, 38-56 | 46, 39-58 | 46, 38-53 | 0.225 |
| Gender |  |  |  | 0.02 |
| Female | 313/423, 74% [69-78%] | 185/265, 70% [64-75%] | 128/158, 81% [74-87%] |  |
| Male | 108/423, 26% [21-30%] | 78/265, 29% [24-35%] | 30/158, 19% [13-26%] |  |
| Non-binary | 2/423, 0.5% [0.08-1.9%] | 2/265, 0.8% [0.13-3.0%] | 0/158, 0% [0.00-3.0%] |  |
| Race and ethnicity |  |  |  | 0.46 |
| Asian | 11/423, 2.6% [1.4-4.7%] | 9/265, 3.4% [1.7-6.6%] | 2/158, 1.3% [0.22-5.0%] |  |
| Black | 8/423, 1.9% [0.88-3.8%] | 6/265, 2.3% [0.92-5.1%] | 2/158, 1.3% [0.22-5.0%] |  |
| Latino | 15/423, 3.5% [2.1-5.9%] | 7/265, 2.6% [1.2-5.6%] | 8/158, 5.1% [2.4-10%] |  |
| Multiracial or Other | 21/423, 5.0% [3.2-7.6%] | 13/265, 4.9% [2.7-8.4%] | 8/158, 5.1% [2.4-10%] |  |
| Non-Hispanic White | 368/423, 87% [83-90%] | 230/265, 87% [82-91%] | 138/158, 87% [81-92%] |  |
| Country of residence |  |  |  | 0.81 |
| Canada | 11/423, 2.6% [1.4-4.7%] | 7/265, 2.6% [1.2-5.6%] | 4/158, 2.5% [0.81-6.8%] |  |
| Germany | 5/423, 1.2% [0.44-2.9%] | 2/265, 0.8% [0.13-3.0%] | 3/158, 1.9% [0.49-5.9%] |  |
| United Kingdom | 5/423, 1.2% [0.44-2.9%] | 3/265, 1.1% [0.29-3.5%] | 2/158, 1.3% [0.22-5.0%] |  |
| U.S. | 391/423, 92% [89-95%] | 247/265, 93% [89-96%] | 144/158, 91% [85-95%] |  |
| Other countries | 11/423, 2.6% [1.4-4.7%] | 6/265, 2.3% [0.92-5.1%] | 5/158, 3.2% [1.2-7.6%] |  |
| Marital status |  |  |  | 0.96 |
| Divorced | 42/398, 11% [7.8-14%] | 27/253, 11% [7.3-15%] | 15/145, 10% [6.1-17%] |  |
| Married or civil union | 237/398,<br>60% [55-64%] | 151/253, 60%<br>[53-66%] | 86/145, 59%<br>[51-67%] |  |
| Never married | 109/398,<br>27% [23-32%] | 69/253, 27%<br>[22-33%] | 40/145, 28%<br>[21-36%] |  |
| Separated | 6/398, 1.5%<br>[0.61-3.4%] | 3/253, 1.2%<br>[0.31-3.7%] | 3/145, 2.1%<br>[0.54-6.4%] |  |
| Widowed | 4/398, 1.0%<br>[0.32-2.7%] | 3/253, 1.2%<br>[0.31-3.7%] | 1/145, 0.7%<br>[0.04-4.4%] |  |
| Missing data | 25 | 12 | 13 |  |
| Employed pre-pandemic | 338/396,<br>85% [81-89%] | 213/252, 85%<br>[79-89%] | 125/144, 87%<br>[80-92%] | 0.54 |
| Missing data | 27 | 13 | 14 |  |
| Pre-pandemic annual household income |  |  |  | 0.68 |
| \$10,000 to \$35,000 | 20/396, 5.1%<br>[3.2-7.8%] | 12/252, 4.8%<br>[2.6-8.4%] | 8/144, 5.6%<br>[2.6-11%] | |
| \$35,000 to less than \$50,000 | 29/396, 7.3%<br>[5.0-10%] | 15/252, 6.0%<br>[3.5-9.8%] | 14/144, 9.7%<br>[5.6-16%] | |
| \$50,000 to less than \$75,000 | 38/396, 9.6%<br>[7.0-13%] | 25/252, 9.9%<br>[6.6-14%] | 13/144, 9.0%<br>[5.1-15%] | |
| \$75,000 or more | 274/396,<br>69% [64-74%] | 176/252, 70%<br>[64-75%] | 98/144, 68%<br>[60-75%] | |
| Less than \$10,000 | 4/396, 1.0%<br>[0.32-2.7%] | 2/252, 0.8%<br>[0.14-3.1%] | 2/144, 1.4%<br>[0.24-5.4%] | |
| Prefer not to answer | 31/396, 7.8%<br>[5.5-11%] | 22/252, 8.7%<br>[5.7-13%] | 9/144, 6.2%<br>[3.1-12%] |  |
| Missing data | 27 | 13 | 14 |  |
| Index SARS-CoV-2 infection time period |  |  |  | 0.006 |
| Pre-Delta | 171/373,<br>46% [41-51%] | 97/233, 42%<br>[35-48%] | 74/140, 53%<br>[44-61%] |  |
| Delta | 48/373, 13%<br>[9.7-17%] | 27/233, 12%<br>[7.9-17%] | 21/140, 15%<br>[9.7-22%] |  |
| Omicron | 110/373,<br>29% [25-34%] | 72/233, 31%<br>[25-37%] | 38/140, 27%<br>[20-35%] |  |
| Post-Omicron | 44/373, 12%<br>[8.8-16%] | 37/233, 16%<br>[12-21%] | 7/140, 5.0%<br>[2.2-10%] |  |
| Missing data | 50 | 32 | 18 |  |
| Hospitalized for COVID-related conditions | 40/423, 9.5%<br>[6.9-13%] | 20/265, 7.5%<br>[4.8-12%] | 20/158, 13%<br>[8.1-19%] | 0.08 |
| Number of weeks between infection and completing the symptom survey | 63, 32-111 | 54, 26-99 | 74, 38-118 | 0.003 |
| Missing data | 50 | 32 | 18 |  |
| Financial difficulties caused by the pandemic |  |  |  | <0.001 |
| Not at all | 125/396, 32% [27-36%] | 94/252, 37% [31-44%] | 31/144, 22% [15-29%] |  |
| A little | 146/396, 37% [32-42%] | 95/252, 38% [32-44%] | 51/144, 35% [28-44%] |  |
| Quite a bit | 66/396, 17% [13-21%] | 36/252, 14% [10-19%] | 30/144, 21% [15-29%] |  |
| Very much | 59/396, 15% [12-19%] | 27/252, 11% [7.3-15%] | 32/144, 22% [16-30%] |  |
| Missing data | 27 | 13 | 14 |  |
| Do not have health insurance | 15/423, 3.5% [2.1-5.9%] | 9/265, 3.4% [1.7-6.6%] | 6/158, 3.8% [1.6-8.5%] | 0.83 |
| Social support (someone around to help you if you need it) |  |  |  | 0.77 |
| Never | 24/398, 6.0% [4.0-9.0%] | 16/253, 6.3% [3.8-10%] | 8/145, 5.5% [2.6-11%] |  |
| Rarely | 31/398, 7.8% [5.4-11%] | 22/253, 8.7% [5.7-13%] | 9/145, 6.2% [3.1-12%] |  |
| Sometimes | 72/398, 18% [15-22%] | 42/253, 17% [12-22%] | 30/145, 21% [15-28%] |  |
| Usually | 148/398, 37% [32-42%] | 93/253, 37% [31-43%] | 55/145, 38% [30-46%] |  |
| Always | 123/398, 31% [26-36%] | 80/253, 32% [26-38%] | 43/145, 30% [23-38%] |  |
| Missing data | 25 | 12 | 13 |  |
| Social isolation (how often do you feel isolated from others?) |  |  |  | 0.04 |
| Hardly ever or never | 85/395, 22% [18-26%] | 64/251, 25% [20-31%] | 21/144, 15% [9.5-22%] |  |
| Some of the time | 155/395, 39% [34-44%] | 94/251, 37% [32-44%] | 61/144, 42% [34-51%] |  |
| Often | 155/395, 39% [34-44%] | 93/251, 37% [31-43%] | 62/144, 43% [35-52%] |  |
| Missing data | 28 | 14 | 14 |  |
| Housing |  |  |  | <0.001 |
| I do not have a steady place to live | 4/398, 1.0%<br>[0.32-2.7%] | 0/253, 0%<br>[0.00-1.9%] | 4/145, 2.8%<br>[0.89-7.4%] |  |
| I have a place to live today, but I am worried about losing it in the future | 47/398, 12%<br>[8.9-15%] | 21/253, 8.3%<br>[5.3-13%] | 26/145, 18%<br>[12-25%] |  |
| I have a steady place to live | 347/398, 87%<br>[83-90%] | 232/253, 92%<br>[87-95%] | 115/145, 79%<br>[72-85%] |  |
| Missing data | 25 | 12 | 13 |  |
| Abbreviations: CI, confidence interval |  |  |  |  |
| <sup>1</sup> Wilcoxon rank sum test; Fisher's exact test; Pearson's Chi-squared test |  |  |  |  |

### Pre-pandemic comorbidities

Overall, among participants, the most common self-reported pre-pandemic comorbidities were anxiety disorders (30%), depressive disorders (29%), and gastrointestinal issues, including irritable bowel syndrome and acid reflux (24%). Participants with and without internal tremors had similar rates of all self-reported pre-pandemic comorbidities (eTable 1).

### SARS-CoV-2 infection characteristics and post-COVID socioeconomic characteristics

Overall, the most common period for index SARS-CoV-2 infection was during the Pre-Delta wave (46%); 9.5% of participants were hospitalized due to COVID-related conditions (Table 1, eFigure 1). Participants with internal tremors were significantly more likely to report their index infection during the Pre-Delta wave (53% [95% CI, 44-61] vs. 42% [35-48], P = 0.006) but were not significantly different in hospitalization rates due to COVID-related conditions. Participants with internal tremors had a significantly longer duration between their initial infections and the date of completing LISTEN’s symptom survey (median: 74 weeks [IQR, 38-118] vs. 54 weeks [26-99], P = 0.003).

Participants with and without internal tremors had no significant differences in their health insurance status and level of social support when completing the surveys (P > 0.05; Table 1). Participants with internal tremors were significantly more likely to report having financial difficulties caused by the pandemic (very much financial difficulties, 22% [95% CI, 16-30] vs. 11% [7.3-15], P < 0.001), often feeling socially isolated (43% [95% CI, 35-52] vs. 37% [31-43], P = 0.04), and housing insecurity (worried about losing housing, 18% [95% CI, 12-25] vs. 8.3% [5.3-13], P < 0.001).

### Health status

Participants with internal tremors reported significantly worse health as measured by EQ-VAS (median: 40 points [IQR, 30-60] vs. 50 points [35-62], P = 0.007) compared with those with no internal tremors (Table 2). When asked to rate their symptom severity on their worst days using a visual sliding scale ranging from 0 to 100, with 0 being a trivial illness and 100 being unbearable, participants with internal tremors reported greater symptom severity compared with those with no internal tremors (median: 80 points [IQR 73–90] vs. 73 points [60–82], respectively, P < 0.001; Table 2). In both groups, the time period of the index SARS-CoV-2 infection was not significantly associated with EQ-VAS (eTable 2).

**Table 2.** Health status, symptom severity, and RECOVER score.

|  | **Overall,**<br>**N = 423** | **No internal**<br>**tremors, N =**<br>**265** | **Has internal**<br>**tremors, N =**<br>**158** | **p-**<br>**value<sup>1</sup>** |
| --- | --- | --- | --- | --- |
| Euro-QoL visual analogue scale (0-100, 100 means best), median (IQR) | 49 (32, 61) | 50 (35, 62) | 40 (30, 60) | 0.007 |
| Symptom severity on worst days (0-100, 100 means unbearable), median (IQR) | 79 (65, 86) | 73 (60, 82) | 80 (73, 90) | <0.001 |
| Missing data | 24 | 11 | 13 |  |
| RECOVER composite symptom score, median (IQR) | 16 (12, 23) | 15 (11, 21) | 20 (14, 26) | <0.001 |
| RECOVER composite symptom score greater than 12, No. (%) [95% CI] | 332 (78) [74-82] | 191 (72) [66-77] | 141 (89) [83-93] | <0.001 |
| RECOVER composite symptom score (exclude symptoms with severity criteria), median (IQR) | 12 (8, 19) | 11 (8, 17) | 15 (10, 22) | <0.001 |
| RECOVER composite symptom score greater than 12 (exclude symptoms with severity criteria), No. (%) [95% CI] | 216 (51) [46-56] | 111 (42) [36-48] | 105 (66) [58-74] | <0.001 |
| Abbreviations: CI = Confidence Interval, IQR=interquartile range, RECOVER=Researching COVID to Enhance Recovery. |  |  |  |  |
| Symptom severity assessed by the question, “How bad your long COVID or other symptoms are (0 to 100) on your worst days?” See full question phrasing in eMethods 1. |  |  |  |  |
| <sup>1</sup> Wilcoxon rank sum test; Pearson’s Chi-squared test |  |  |  |  |

### New-onset conditions

Overall, the most common new-onset conditions among all participants were postural orthostatic tachycardia syndrome or other dysautonomia (25%), gastrointestinal issues (15%), and neurologic conditions (13%) (eTable 3). Compared with participants without internal tremors, significantly greater proportions of participants with internal tremors reported new-onset mast cell disorders (11% [95% CI, 7.1-18] vs. 2.6% [1.2-5.6]), neurologic conditions (22% [95% CI, 16-29] vs. 8.3% [5.4-12]), anxiety disorders (20% [95% CI, 14-27] vs. 8.7% [5.7-13]), and trauma- and stressor-related disorders (12% [95% CI, 7.6-18] vs. 3.4% [1.7-6.6] (Bonferroni-adjusted P < 0.05 for each; Figure 2, eTable 3).

**Figure 2.**
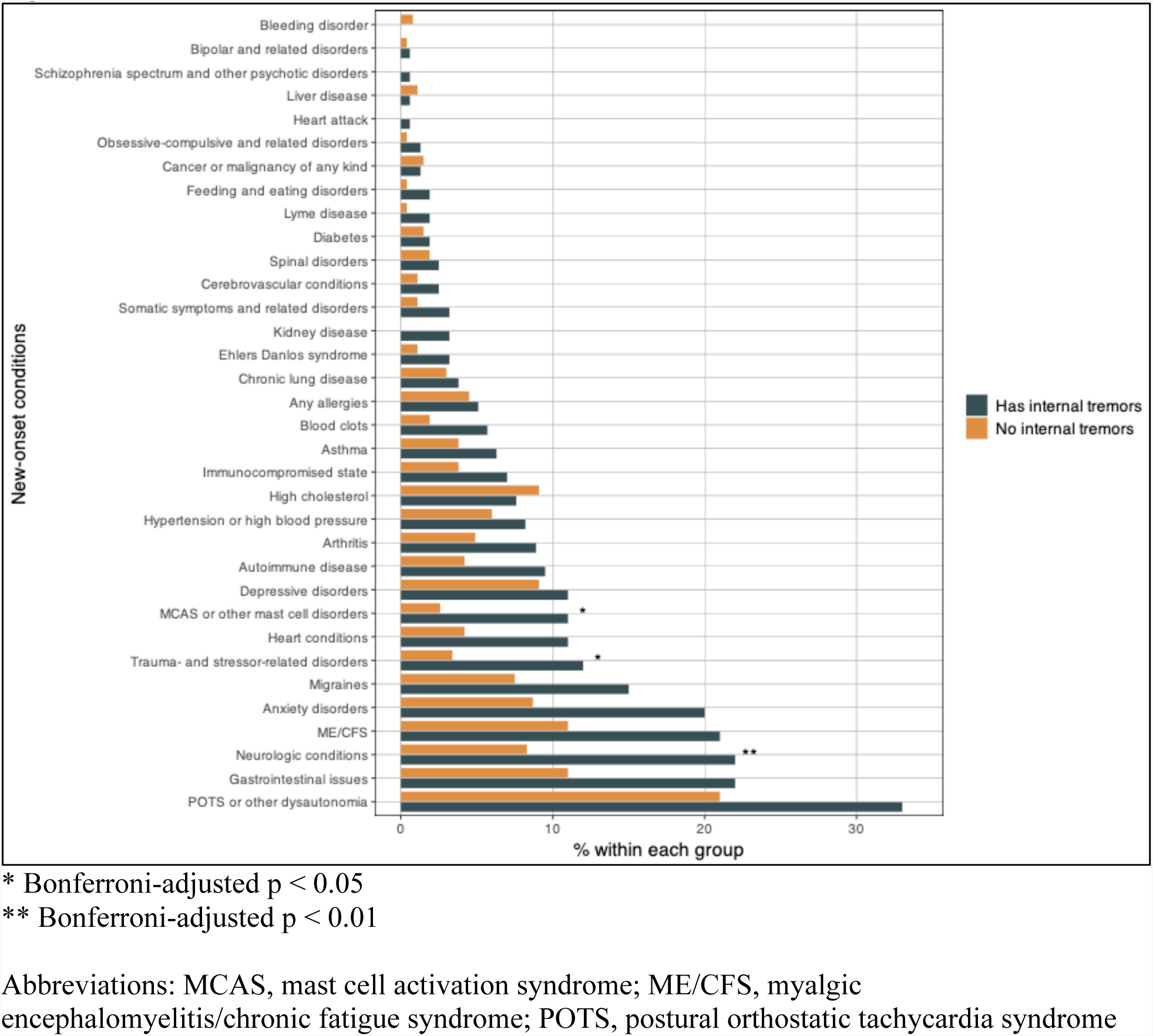
New-onset conditions.

### RECOVER score

Overall, the most common long COVID symptoms reported were excessive fatigue (87%), brain fog (86%), exercise intolerance (79%), trouble falling or staying asleep (72%), and memory problems (70%) (eTable 4). Participants had a median composite symptom score of 16 (IQR, 12-23), based on scoring criteria proposed by the RECOVER Consortium; 78% of participants had a RECOVER score greater than 12 points (Table 2). Compared with participants with no internal tremors, those with internal tremors had significantly higher composite symptom scores (median: 20 [IQR, 14–26] vs. 15 [11, 21], P < 0.001).

### Symptoms differentiating participants experiencing internal tremors compared with those not experiencing internal tremors

We used feature importance in gradient boosted tree machine learning models to identify which symptoms are most important for differentiating between participants who experience internal tremors and those who do not (Figure 3A; eTable 5). We reported the 6 symptom variables that were most important in the selected final gradient boosted model (Figure 3B).

**Figure 3.**
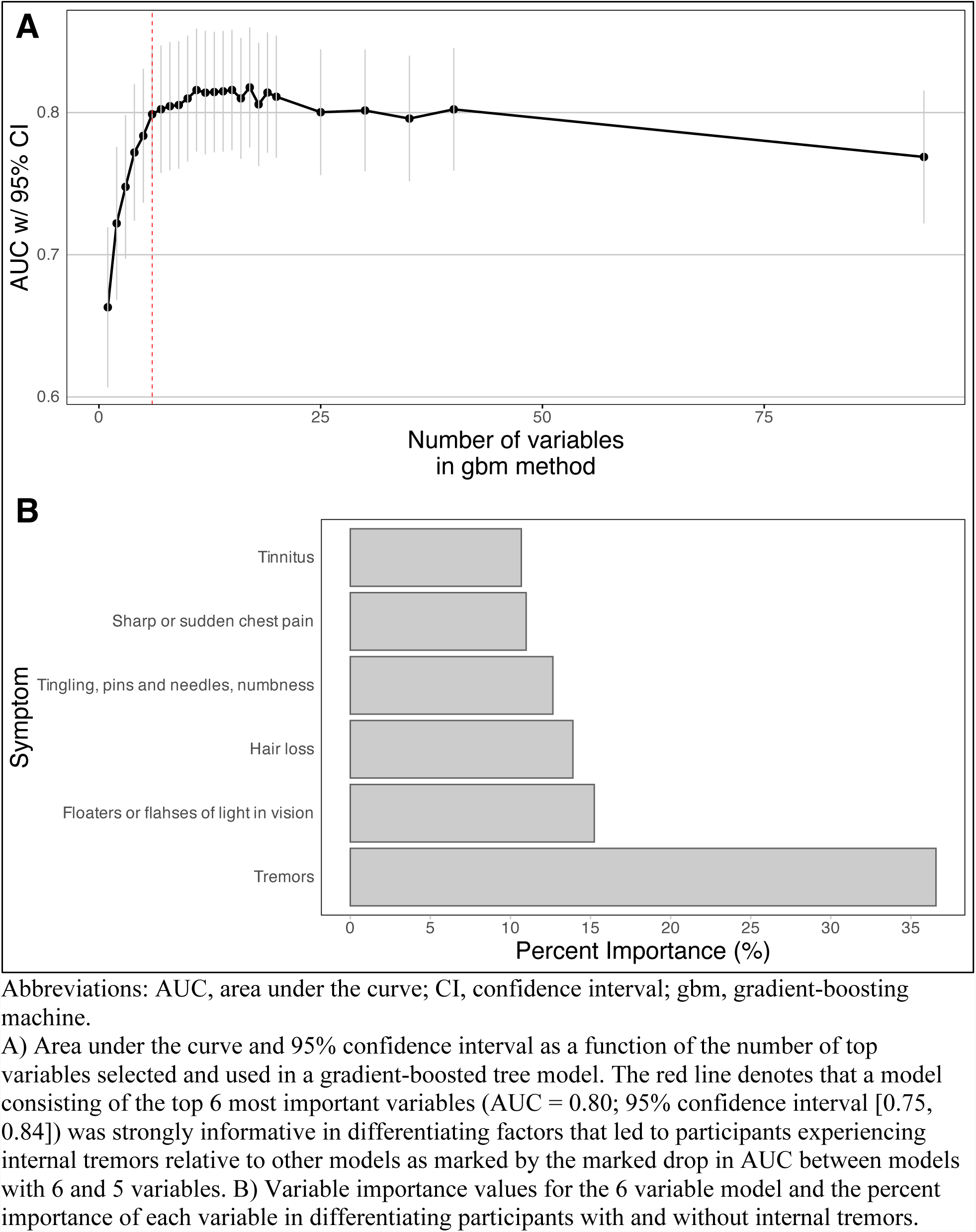
Most important symptoms for differentiating between participants with and without internal tremors.

The same number of variables (n = 6) and overall performance were observed for the selected final models when we repeated our analysis with two other methods: XGBoost Gain and XGBoost Shap (eFigure 2). The final models in each of the three methods were compared and the selected variables and their relative rankings were concordant (eFigure 3A). The Pearson correlation of the non-zero variable importance values between each of the methods was between 0.97 to 0.99, all with p-value < 1.3e-11, further suggesting that the results of the modeling approach were robust.

Participants with internal tremors had significantly higher rates for all 6 of the most important symptoms, compared to participants without internal tremors: tremors or shakiness (65% [95% CI, 57-72] vs. 22% [18-28]), floaters or flashes of light in vision (47% [95% CI, 39-55] vs. 16% [12-21]), hair loss (56% [95% CI, 48-64] vs. 30% [24-36]), tingling, pins and needles, and numbness (72% [95% CI, 64-78]) vs. 41% [35-47]), sharp or sudden chest pain (47% [95% CI, 39-55] vs. 22% [17-27]), and tinnitus or humming in ears (62% [95% CI, 54-70] vs. 36% [30-42]) (Bonferroni-adjusted P < 0.05 for each; eTable 5).

## DISCUSSION

In a cross-sectional study of people with long COVID, we found internal tremors were a common symptom, affecting 37% of participants. Participants with internal tremors were more likely to be female but had otherwise similar demographic characteristics compared to those without internal tremors. Importantly, although the two groups had similar pre-pandemic comorbidities, participants with internal tremors had, at the time of the survey, worse EQ-VAS health status, higher RECOVER composite symptoms scores, and higher rates of new-onset conditions for mast cell disorders, neurologic disorders, anxiety disorders, and trauma- and stressor-related disorders. Symptoms important for differentiating participants with and without internal tremors included neurologic symptoms such as tremors or shakiness, floaters or flashes of light in vision, tinnitus or humming in ears, and tingling, pins and needles, and numbness. Socioeconomically, participants with internal tremors had more financial difficulties caused by the pandemic, social isolation, and housing insecurity.

To the best of our knowledge, only four prior studies have described people with internal tremors;^3–5, 18^ no quantitative study of people with long COVID has fully described symptoms of internal tremors. Two recent studies based on large prospective cohorts of people with long COVID did not report on symptoms of internal tremors.^7, 11^ Our study included questions about internal tremors based on suggestions from participants. Our findings are consistent with and extend prior findings by showing that compared with others with long COVID, participants with internal tremors had worse quality of life than others with long COVID and had significantly higher rates of neurologic symptoms.

Rates of new-onset dysautonomia were higher among participants with internal tremors, with statistical significance before but not after correction. The mechanism of dysautonomia among patients with Parkinson’s disease may be related to organ-selective sympathetic denervation,^19^ but the pathophysiologic links among dysautonomia, long COVID, and internal tremors have not been established and should be explored in further studies.

Our findings add nuance to the hypothesized associations between myalgic encephalomyelitis/chronic fatigue syndrome and long COVID, which are often discussed together due to overlapping symptomatology and infectious etiopathogenesis.^20–22^ Although greater than 70% of participants in this study reported excessive fatigue or exercise intolerance, these hallmark symptoms of myalgic encephalomyelitis/chronic fatigue syndrome were not significantly associated with internal tremors. Our findings are consistent with prior observations of long COVID as a heterogeneous condition with many phenotypes,^23^ which may be driven by several mechanisms.^24^

### Study limitations

This study has several limitations. Study participants were recruited from an online long COVID community and should not be considered representative. Rather, the study provides an opportunity to describe the characteristics of people within the online community who do and do not have internal tremors and vibrations. In addition, conditions and symptoms are based on self-reporting using a pre-specified checklist, which may be susceptible to recall bias with unclear directionality for under versus overreporting, as well as inaccuracies such as misdiagnoses due to uncertainties surrounding long COVID evaluation and management during early stages of the pandemic.

### Conclusions

Internal tremors and vibrations are common symptoms among people with long COVID. People with these symptoms had pre-infection characteristics like those of others with long COVID, but compared with others who had long COVID, they had worse EQ-VAS health status and higher rates of financial difficulties and housing insecurity, and higher self-reported rates of new-onset conditions of mast cell disorders, neurologic conditions, anxiety, and trauma- and stressor-related conditions. Individuals with long COVID symptoms of internal tremors may experience a particularly severe phenotype of long COVID. Clinicians should be aware of internal tremors and vibrations as a long COVID symptom. Further research is needed to clarify the pathophysiology of internal tremors and vibrations and identify potential treatment targets.

## Data Availability

Code and deidentified data to reproduce machine learning analyses in the present study are publicly available. Other deidentified data in the present study are available upon reasonable request to the authors.

https://github.com/aditharun/tremors-ml

## ACKNOWLEDGMENTS

We thank Shaun Barcavage, FNP-BC, James Curry, MD, and Kelley Royer, RN for their feedback and for sharing their lived experiences with the writing team, and Maria Johnson for her editorial contributions.

## FUNDING

This study was in part funded by the Howard Hughes Medical Institute Collaborative COVID-19 Initiative, and grants from the National Institute of Allergy and Infectious Diseases (R01AI157488 to Akiko Iwasaki).

## CONFLICTS OF INTEREST

Dr. Harlan Krumholz and his spouse are co-founders of, and have equity in, Hugo Health, the personalized health data-platform company that developed the Hugo Health Kindred platform. Dr. Krumholz is a scientific advisor to, and his spouse is an officer with, Hugo Health. His involvement in this study is overseen by the Yale Conflict of Interest Committee. In the past three years, Harlan Krumholz received expenses and/or personal fees from UnitedHealth, Element Science, Eyedentifeye, and F-Prime. He is a co-founder of Refactor Health and is associated with contracts, through Yale New Haven Hospital, from the Centers for Medicare & Medicaid Services and through Yale University from the Food and Drug Administration, Johnson & Johnson, Google, and Pfizer.

Dr. Rohan Khera is an Associate Editor of JAMA. He receives support from the National Heart, Lung, and Blood Institute of the National Institutes of Health (under award K23HL153775) and the Doris Duke Charitable Foundation (under award, 2022060). He also receives research support, through Yale, from Bristol-Myers Squibb and Novo Nordisk. He is a coinventor of U.S. Provisional Patent Applications 63/177,117, 63/428,569, 63/346,610, and 63/484,426. He is a co-founder of Evidence2Health, a precision health platform to improve evidence-based cardiovascular care.

Dr. Akiko Iwasaki is a co-founder and consults for RIGImmune, Xanadu Bio and PanV, she consults for Paratus Sciences, InvisiShield Technologies, and is a member of the Board of Directors of Roche Holding Ltd.

## Notes

### Author Declarations

The Yale University Institutional Review Board gave ethical approval for this work.

### Summary of Updates

The Methods, Results, Discussion, figures, tables, and supplemental files were updated to reflect new analyses (machine learning models).

