## Supplement for "Internal tremors and vibrations in long COVID: a cross-sectional study"

Supplement - Table of Contents

**eFigure 1. Distribution of SARS-CoV-2 index infection dates**

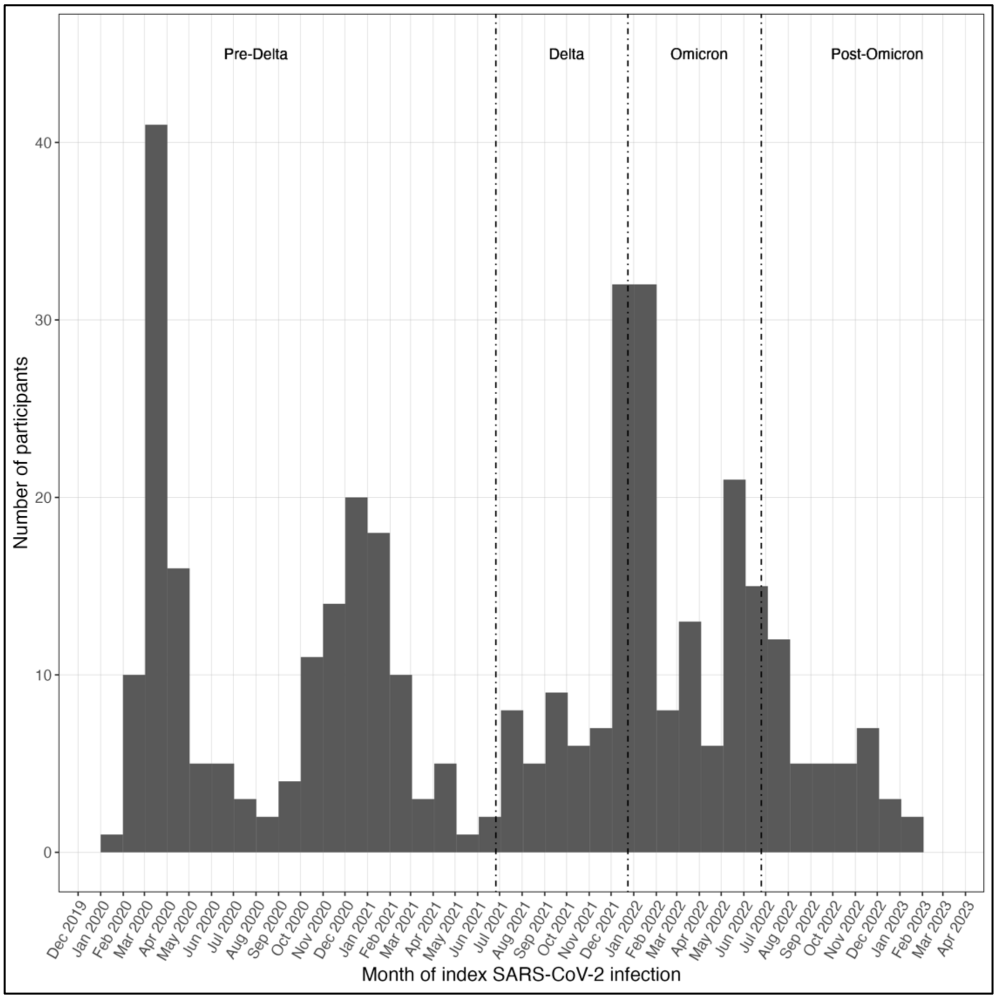

Self-reported time of index SARS-CoV-2 infection was categorized as Pre-Delta (before 26 June 2021), Delta (26 June 2021–24 December 2021), Omicron (25 December 2021–25 June 2022), and Post-Omicron (after 25 June 2022), consistent with time period definitions associated with dominant variants of SARS-CoV-2.
Reference: Gottlieb M, Wang RC, Yu H, et al. Severe Fatigue and Persistent Symptoms at 3 Months Following Severe Acute Respiratory Syndrome Coronavirus 2 Infections During the Pre-Delta, Delta, and Omicron Time Periods: A Multicenter Prospective Cohort Study. *Clin Infect Dis*. 2023;76(11):1930-1941. doi:10.1093/cid/ciad045

**eFigure 2. XGBoost model results for symptoms strongly associated with internal tremors**

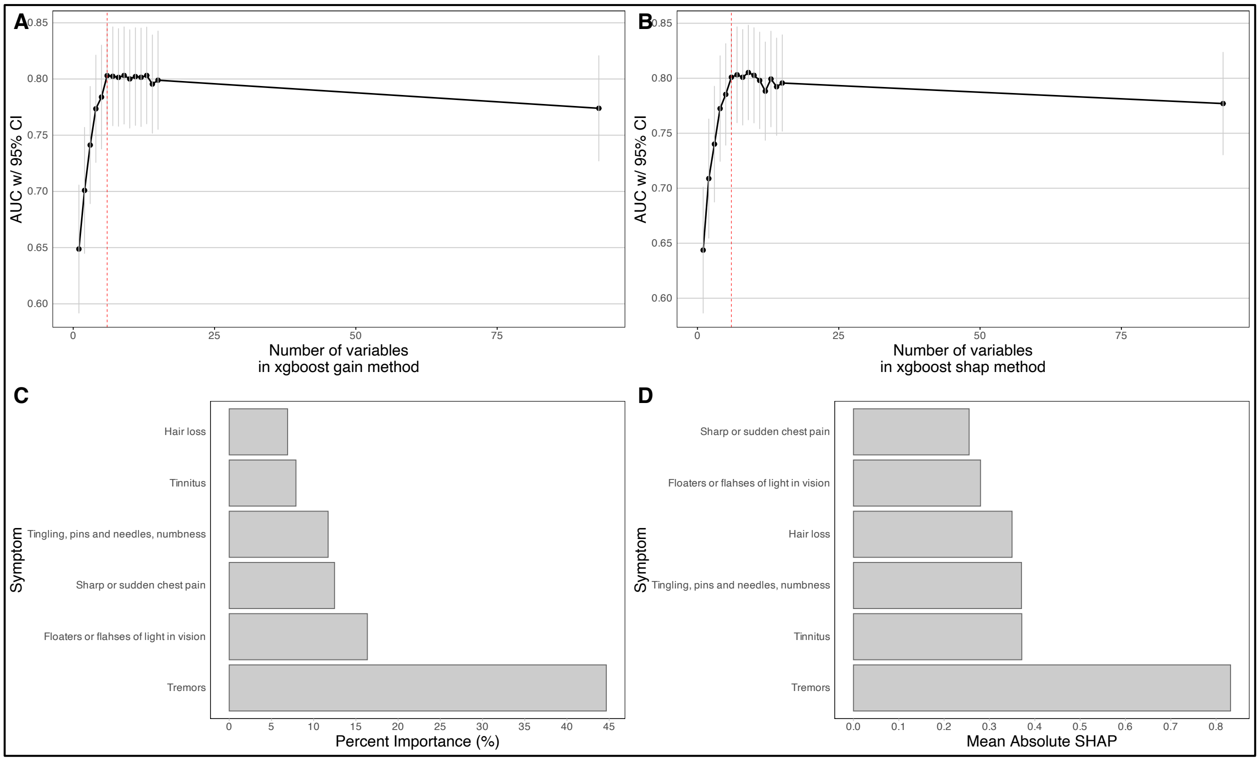

Abbreviations: AUC, area under the curve; CI, confidence interval; SHAP, Shapley value.

A) Model performance as a function of the number of top variables used in XGBoost model with variable importance computed by relative gain. Red line denotes model with 6 variables selected (AUC = 0.80; 95% CI [0.76, 0.85]). B) Model performance as a function of the number of top variables used in XGBoost model with variable importance computed by mean absolute shapley values. Red line denotes model with 6 variables selected (AUC = 0.80; 95% CI [0.76, 0.85]). C) Symptoms' importance in differentiating internal tremor symptom status for XGBoost gain model selected in panel A. D) Symptoms' importance in differentiating internal tremor symptom status for XGBoost shap model selected in panel B.

**eFigure 3. Comparison between symptoms associated with internal tremors across machine learning methods**

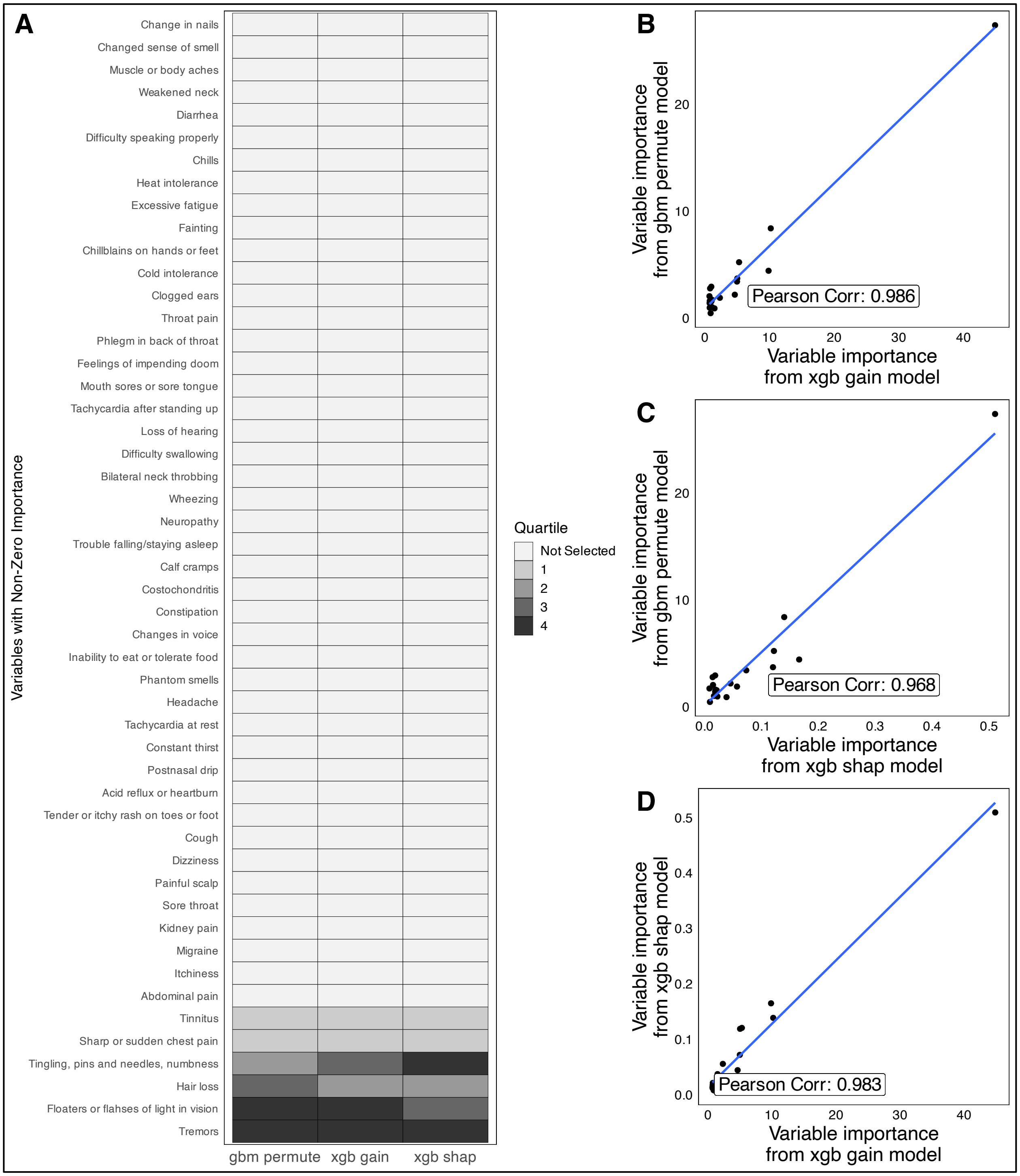

Abbreviations: gbm permute, gradient-boosting machine using a permutation-based approach; xgb gain, XGBoost tree machine learning model with gain in accuracy metric; xgb shap, XGBoost tree machine learning model with Shapley value.

A) Comparison between the selected final models for each of the three methods. All variables with non-zero importance are shown and for each of the three models, each variable is classified by their quartile rank in the final model or is not selected. The selected variables are highly concordant across models suggesting that the variables selected are truly important in differentiating patients with and without internal tremors.

B, C, D) Correlation between the variable importance metrics for each of the three methods for all variables with non-zero importances. B) Correlation between XGBoost gain and GBM permute is 0.99 with $p < 1.19e-14$. C) Correlation between XGBoost shap and GBM permute is 0.97 with $p < 1.3e-11$. D) Correlation between XGBoost gain and XGBoost shap is 0.98 with $p < 2.2e-16$.

**eTable 1. Pre-pandemic comorbidities**

| | **Overall**, N = 423 | **No internal tremors**, N = 265 | **Has internal tremors**, N = 158 | **p-value**[1] | **Adjusted p-value**[2] |
|---|---|---|---|---|---|
| | **n/N, % [95% CI]** | **n/N, % [95% CI]** | **n/N, % [95% CI]** | | |
| Any allergies | 208/423, 49% [44-54%] | 141/265, 53% [47-59%] | 67/158, 42% [35-51%] | 0.032 | >0.999 |
| Arthritis (including rheumatoid arthritis, gout, lupus, or fibromyalgia) | 67/423, 16% [13-20%] | 46/265, 17% [13-23%] | 21/158, 13% [8.6-20%] | 0.268 | >0.999 |
| Asthma | 78/423, 18% [15-23%] | 58/265, 22% [17-27%] | 20/158, 13% [8.1-19%] | 0.018 | 0.627 |
| Autoimmune disease (including lupus, scleroderma, etc.) | 50/423, 12% [9.0-15%] | 35/265, 13% [9.5-18%] | 15/158, 9.5% [5.6-15%] | 0.252 | >0.999 |
| Bleeding disorder (including sickle cell disease or thalassemia) | 4/423, 0.9% [0.30-2.6%] | 3/265, 1.1% [0.29-3.5%] | 1/158, 0.6% [0.03-4.0%] | >0.999 | >0.999 |
| Blood clots | 6/423, 1.4% [0.58-3.2%] | 2/265, 0.8% [0.13-3.0%] | 4/158, 2.5% [0.81-6.8%] | 0.202 | >0.999 |
| Cancer or malignancy of any kind | 21/423, 5.0% [3.2-7.6%] | 12/265, 4.5% [2.5-8.0%] | 9/158, 5.7% [2.8-11%] | 0.593 | >0.999 |
| Cerebrovascular conditions affecting blood vessels to or in the brain (including stroke) | 3/423, 0.7% [0.18-2.2%] | 3/265, 1.1% [0.29-3.5%] | 0/158, 0% [0.00-3.0%] | 0.296 | >0.999 |
| Chronic lung disease (emphysema, chronic bronchitis, chronic obstructive pulmonary disease) | 10/423, 2.4% [1.2-4.4%] | 6/265, 2.3% [0.92-5.1%] | 4/158, 2.5% [0.81-6.8%] | >0.999 | >0.999 |
| Diabetes | 11/423, 2.6% | 7/265, 2.6% | 4/158, 2.5% | >0.999 | >0.999 |

| | | | | | |
|---|---|---|---|---|---|
| | [1.4-4.7%] | [1.2-5.6%] | [0.81-6.8%] | | |
| Ehlers Danlos Syndrome (hypermobile joints) | 8/423, 1.9% [0.88-3.8%] | 4/265, 1.5% [0.48-4.1%] | 4/158, 2.5% [0.81-6.8%] | 0.479 | >0.999 |
| Gastrointestinal issues (including IBS or acid reflux) | 103/423, 24% [20-29%] | 67/265, 25% [20-31%] | 36/158, 23% [17-30%] | 0.563 | >0.999 |
| Heart attack, also called myocardial infarction | 3/423, 0.7% [0.18-2.2%] | 2/265, 0.8% [0.13-3.0%] | 1/158, 0.6% [0.03-4.0%] | >0.999 | >0.999 |
| Heart conditions (including coronary artery disease or cardiomyopathies) | 12/423, 2.8% [1.5-5.0%] | 7/265, 2.6% [1.2-5.6%] | 5/158, 3.2% [1.2-7.6%] | 0.768 | >0.999 |
| High cholesterol | 59/423, 14% [11-18%] | 42/265, 16% [12-21%] | 17/158, 11% [6.6-17%] | 0.144 | >0.999 |
| Hypertension or high blood pressure | 50/423, 12% [9.0-15%] | 34/265, 13% [9.2-18%] | 16/158, 10% [6.1-16%] | 0.405 | >0.999 |
| Immunocompromised state | 13/423, 3.1% [1.7-5.3%] | 12/265, 4.5% [2.5-8.0%] | 1/158, 0.6% [0.03-4.0%] | 0.037 | >0.999 |
| Kidney disease | 5/423, 1.2% [0.44-2.9%] | 4/265, 1.5% [0.48-4.1%] | 1/158, 0.6% [0.03-4.0%] | 0.655 | >0.999 |
| Liver disease | 3/423, 0.7% [0.18-2.2%] | 2/265, 0.8% [0.13-3.0%] | 1/158, 0.6% [0.03-4.0%] | >0.999 | >0.999 |
| Lyme disease | 25/423, 5.9% [3.9-8.7%] | 14/265, 5.3% [3.0-8.9%] | 11/158, 7.0% [3.7-12%] | 0.479 | >0.999 |
| MCAS (mast cell activation syndrome) or other mast cell disorders | 4/423, 0.9% [0.30-2.6%] | 1/265, 0.4% [0.02-2.4%] | 3/158, 1.9% [0.49-5.9%] | 0.149 | >0.999 |

| | | | | | |
|---|---|---|---|---|---|
| ME/CFS (myalgic encephalomyelitis/chronic fatigue syndrome) | 16/423, 3.8% [2.3-6.2%] | 10/265, 3.8% [1.9-7.0%] | 6/158, 3.8% [1.6-8.5%] | 0.990 | >0.999 |
| Migraines | 84/423, 20% [16-24%] | 50/265, 19% [14-24%] | 34/158, 22% [16-29%] | 0.509 | >0.999 |
| Neurologic conditions (including seizures, dementia, multiple sclerosis, Parkinson's, neuropathy, small fiber neuropathy, etc.) | 20/423, 4.7% [3.0-7.3%] | 14/265, 5.3% [3.0-8.9%] | 6/158, 3.8% [1.6-8.5%] | 0.486 | >0.999 |
| Postural orthostatic hypotension (POTS) or other dysautonomia | 13/423, 3.1% [1.7-5.3%] | 5/265, 1.9% [0.70-4.6%] | 8/158, 5.1% [2.4-10%] | 0.083 | >0.999 |
| Spinal disorder(s) | 16/423, 3.8% [2.3-6.2%] | 10/265, 3.8% [1.9-7.0%] | 6/158, 3.8% [1.6-8.5%] | 0.990 | >0.999 |
| Tremors/Internal vibrations | 6/423, 1.4% [0.58-3.2%] | 2/265, 0.8% [0.13-3.0%] | 4/158, 2.5% [0.81-6.8%] | 0.202 | >0.999 |
| Depressive disorders | 122/423, 29% [25-33%] | 84/265, 32% [26-38%] | 38/158, 24% [18-32%] | 0.093 | >0.999 |
| Anxiety disorders | 127/423, 30% [26-35%] | 83/265, 31% [26-37%] | 44/158, 28% [21-36%] | 0.451 | >0.999 |
| Schizophrenia spectrum and other psychotic disorders | 1/423, 0.2% [0.01-1.5%] | 1/265, 0.4% [0.02-2.4%] | 0/158, 0% [0.00-3.0%] | >0.999 | >0.999 |
| Bipolar and related disorders | 13/423, 3.1% [1.7-5.3%] | 6/265, 2.3% [0.92-5.1%] | 7/158, 4.4% [2.0-9.3%] | 0.249 | >0.999 |
| Obsessive-compulsive and related disorders | 15/423, 3.5% [2.1-5.9%] | 7/265, 2.6% [1.2-5.6%] | 8/158, 5.1% [2.4-10%] | 0.193 | >0.999 |

| | | | | | |
|---|---|---|---|---|---|
| Trauma- and stressor-related disorders | 46/423, 11% [8.1-14%] | 27/265, 10% [6.9-15%] | 19/158, 12% [7.6-18%] | 0.557 | >0.999 |
| Feeding and eating disorders | 20/423, 4.7% [3.0-7.3%] | 13/265, 4.9% [2.7-8.4%] | 7/158, 4.4% [2.0-9.3%] | 0.824 | >0.999 |
| Somatic symptoms (excessive thoughts, feelings and behaviors relating to physical symptoms) and related disorders | 7/423, 1.7% [0.73-3.5%] | 5/265, 1.9% [0.70-4.6%] | 2/158, 1.3% [0.22-5.0%] | >0.999 | >0.999 |
| [1] Pearson's Chi-squared test; Fisher's exact test | | | | | |
| [2] Bonferroni correction for multiple testing | | | | | |
| CI, confidence interval; IBS, irritable bowel syndrome; POTS, postural orthostatic tachycardia syndrome | | | | | |

**eTable 2. Health status by internal tremors status and time of index SARS-CoV-2 infection**

Excluding n = 50 participants with missing dates of index SARS-CoV-2 infection

| **Overall**, N = 373 | | | | | |
|---|---|---|---|---|---|
| | **Pre-Delta**, N = 171[1] | **Delta**, N = 48[1] | **Omicron**, N = 110[1] | **Post-Omicron**, N = 44[1] | **p-value**[2] |
| Euro-QoL visual analogue scale (0-100) | 41 (32, 60) | 46 (31, 69) | 49 (31, 61) | 50 (34, 60) | 0.914 |
| **No internal tremors,** N = 233 | | | | | |
| | **Pre-Delta**, N = 97[1] | **Delta**, N = 27[1] | **Omicron**, N = 72[1] | **Post-Omicron**, N = 37[1] | **p-value**[2] |
| Euro-QoL visual analogue scale (0-100) | 45 (35, 61) | 59 (38, 69) | 50 (32, 62) | 49 (34, 60) | 0.750 |
| **Has internal tremors,** N = 140 | | | | | |
| | **Pre-Delta**, N = 74[1] | **Delta**, N = 21[1] | **Omicron**, N = 38[1] | **Post-Omicron**, N = 7[1] | **p-value**[2] |
| Euro-QoL visual analogue scale (0-100) | 40 (30, 60) | 35 (31, 55) | 41 (30, 58) | 55 (35, 60) | 0.922 |
| [1] Median (IQR) | | | | | |
| [2] Kruskal-Wallis rank sum test | | | | | |

**eTable 3. New-onset conditions**

| | **Overall**, N = 423 | **No internal tremors**, N = 265 | **Has internal tremors**, N = 158 | **p-value**[1] | **Adjusted p-value**[2] |
|---|---|---|---|---|---|
| | **n/N, % [95% CI]** | **n/N, % [95% CI]** | **n/N, % [95% CI]** | | |
| Any allergies | 20/423, 4.7% [3.0-7.3%] | 12/265, 4.5% [2.5-8.0%] | 8/158, 5.1% [2.4-10%] | 0.802 | >0.999 |
| Arthritis (including rheumatoid arthritis, gout, lupus, or fibromyalgia) | 27/423, 6.4% [4.3-9.3%] | 13/265, 4.9% [2.7-8.4%] | 14/158, 8.9% [5.1-15%] | 0.107 | >0.999 |
| Asthma | 20/423, 4.7% [3.0-7.3%] | 10/265, 3.8% [1.9-7.0%] | 10/158, 6.3% [3.2-12%] | 0.231 | >0.999 |
| Autoimmune disease (including lupus, scleroderma, etc.) | 26/423, 6.1% [4.1-9.0%] | 11/265, 4.2% [2.2-7.5%] | 15/158, 9.5% [5.6-15%] | 0.027 | 0.914 |
| Bleeding disorder (including sickle cell disease or thalassemia) | 2/423, 0.5% [0.08-1.9%] | 2/265, 0.8% [0.13-3.0%] | 0/158, 0% [0.00-3.0%] | 0.531 | >0.999 |
| Blood clots | 14/423, 3.3% [1.9-5.6%] | 5/265, 1.9% [0.70-4.6%] | 9/158, 5.7% [2.8-11%] | 0.034 | >0.999 |
| Cancer or malignancy of any kind | 6/423, 1.4% [0.58-3.2%] | 4/265, 1.5% [0.48-4.1%] | 2/158, 1.3% [0.22-5.0%] | >0.999 | >0.999 |
| Cerebrovascular conditions affecting blood vessels to or in the brain (including stroke) | 7/423, 1.7% [0.73-3.5%] | 3/265, 1.1% [0.29-3.5%] | 4/158, 2.5% [0.81-6.8%] | 0.432 | >0.999 |
| Chronic lung disease (emphysema, chronic bronchitis, chronic obstructive pulmonary disease) | 14/423, 3.3% [1.9-5.6%] | 8/265, 3.0% [1.4-6.1%] | 6/158, 3.8% [1.6-8.5%] | 0.665 | >0.999 |
| Diabetes | 7/423, 1.7% | 4/265, 1.5% | 3/158, 1.9% | 0.716 | >0.999 |

| | | | | | |
|---|---|---|---|---|---|
| | [0.73-3.5%] | [0.48-4.1%] | [0.49-5.9%] | | |
| Ehlers Danlos Syndrome (hypermobile joints) | 8/423, 1.9% [0.88-3.8%] | 3/265, 1.1% [0.29-3.5%] | 5/158, 3.2% [1.2-7.6%] | 0.156 | >0.999 |
| Gastrointestinal issues (including IBS or acid reflux) | 65/423, 15% [12-19%] | 30/265, 11% [7.9-16%] | 35/158, 22% [16-30%] | 0.003 | 0.095 |
| Heart attack, also called myocardial infarction | 1/423, 0.2% [0.01-1.5%] | 0/265, 0% [0.00-1.8%] | 1/158, 0.6% [0.03-4.0%] | 0.374 | >0.999 |
| Heart conditions (including coronary artery disease or cardiomyopathies) | 29/423, 6.9% [4.7-9.8%] | 11/265, 4.2% [2.2-7.5%] | 18/158, 11% [7.1-18%] | 0.004 | 0.148 |
| High cholesterol | 36/423, 8.5% [6.1-12%] | 24/265, 9.1% [6.0-13%] | 12/158, 7.6% [4.2-13%] | 0.602 | >0.999 |
| Hypertension or high blood pressure | 29/423, 6.9% [4.7-9.8%] | 16/265, 6.0% [3.6-9.8%] | 13/158, 8.2% [4.6-14%] | 0.389 | >0.999 |
| Immunocompromised state | 21/423, 5.0% [3.2-7.6%] | 10/265, 3.8% [1.9-7.0%] | 11/158, 7.0% [3.7-12%] | 0.144 | >0.999 |
| Kidney disease | 5/423, 1.2% [0.44-2.9%] | 0/265, 0% [0.00-1.8%] | 5/158, 3.2% [1.2-7.6%] | 0.007 | 0.237 |
| Liver disease | 4/423, 0.9% [0.30-2.6%] | 3/265, 1.1% [0.29-3.5%] | 1/158, 0.6% [0.03-4.0%] | >0.999 | >0.999 |
| Lyme disease | 4/423, 0.9% [0.30-2.6%] | 1/265, 0.4% [0.02-2.4%] | 3/158, 1.9% [0.49-5.9%] | 0.149 | >0.999 |
| MCAS (mast cell activation syndrome) or other mast cell disorders | 25/423, 5.9% [3.9-8.7%] | 7/265, 2.6% [1.2-5.6%] | 18/158, 11% [7.1-18%] | <0.001 | 0.008 |

| | | | | | |
|---|---|---|---|---|---|
| ME/CFS (myalgic encephalomyelitis/chronic fatigue syndrome) | 62/423, 15% [11-18%] | 29/265, 11% [7.6-15%] | 33/158, 21% [15-28%] | 0.005 | 0.175 |
| Migraines | 43/423, 10% [7.5-14%] | 20/265, 7.5% [4.8-12%] | 23/158, 15% [9.6-21%] | 0.021 | 0.714 |
| Neurologic conditions (including seizures, dementia, multiple sclerosis, Parkinson's, neuropathy, small fiber neuropathy, etc.) | 56/423, 13% [10-17%] | 22/265, 8.3% [5.4-12%] | 34/158, 22% [16-29%] | <0.001 | 0.004 |
| Postural orthostatic hypotension (POTS) or other dysautonomia | 107/423, 25% [21-30%] | 55/265, 21% [16-26%] | 52/158, 33% [26-41%] | 0.005 | 0.184 |
| Spinal disorder(s) | 9/423, 2.1% [1.0-4.1%] | 5/265, 1.9% [0.70-4.6%] | 4/158, 2.5% [0.81-6.8%] | 0.733 | >0.999 |
| Depressive disorders | 42/423, 9.9% [7.3-13%] | 24/265, 9.1% [6.0-13%] | 18/158, 11% [7.1-18%] | 0.437 | >0.999 |
| Anxiety disorders | 54/423, 13% [9.8-16%] | 23/265, 8.7% [5.7-13%] | 31/158, 20% [14-27%] | 0.001 | 0.038 |
| Schizophrenia spectrum and other psychotic disorders | 1/423, 0.2% [0.01-1.5%] | 0/265, 0% [0.00-1.8%] | 1/158, 0.6% [0.03-4.0%] | 0.374 | >0.999 |
| Bipolar and related disorders | 2/423, 0.5% [0.08-1.9%] | 1/265, 0.4% [0.02-2.4%] | 1/158, 0.6% [0.03-4.0%] | >0.999 | >0.999 |
| Obsessive-compulsive and related disorders | 3/423, 0.7% [0.18-2.2%] | 1/265, 0.4% [0.02-2.4%] | 2/158, 1.3% [0.22-5.0%] | 0.559 | >0.999 |
| Trauma- and stressor-related disorders | 28/423, 6.6% [4.5-9.5%] | 9/265, 3.4% [1.7-6.6%] | 19/158, 12% [7.6-18%] | <0.001 | 0.019 |

| | | | | | |
|---|---|---|---|---|---|
| Feeding and eating disorders | 4/423, 0.9% [0.30-2.6%] | 1/265, 0.4% [0.02-2.4%] | 3/158, 1.9% [0.49-5.9%] | 0.149 | >0.999 |
| Somatic symptoms (excessive thoughts, feelings and behaviors relating to physical symptoms) and related disorders | 8/423, 1.9% [0.88-3.8%] | 3/265, 1.1% [0.29-3.5%] | 5/158, 3.2% [1.2-7.6%] | 0.156 | >0.999 |
| [1] Pearson's Chi-squared test; Fisher's exact test | | | | | |
| [2] Bonferroni correction for multiple testing | | | | | |
| CI, confidence interval; IBS, irritable bowel syndrome; POTS, postural orthostatic tachycardia syndrome | | | | | |

**eTable 4. Long COVID symptoms**

| | **Overall**, N = 423 | **No internal tremors**, N = 265 | **Has internal tremors**, N = 158 | **p-value**[1] | **Adjusted p-value**[2] |
|---|---|---|---|---|---|
| | **n/N, % [95% CI]** | **n/N, % [95% CI]** | **n/N, % [95% CI]** | | |
| Anxiety | 185/423, 44% [39-49%] | 97/265, 37% [31-43%] | 88/158, 56% [48-64%] | <0.001 | **0.012** |
| Confusion | 166/423, 39% [35-44%] | 91/265, 34% [29-40%] | 75/158, 47% [40-56%] | 0.007 | 0.710 |
| Brain fog; difficulty concentrating or focusing | 362/423, 86% [82-89%] | 223/265, 84% [79-88%] | 139/158, 88% [82-92%] | 0.279 | >0.999 |
| Feelings of impending doom | 105/423, 25% [21-29%] | 50/265, 19% [14-24%] | 55/158, 35% [28-43%] | <0.001 | **0.023** |
| Memory problems | 297/423, 70% [66-74%] | 175/265, 66% [60-72%] | 122/158, 77% [70-83%] | 0.015 | >0.999 |
| Difficulty speaking properly | 195/423, 46% [41-51%] | 106/265, 40% [34-46%] | 89/158, 56% [48-64%] | 0.001 | 0.106 |
| Suicidal thoughts | 60/423, 14% [11-18%] | 27/265, 10% [6.9-15%] | 33/158, 21% [15-28%] | 0.002 | 0.217 |
| Abnormally low temperature | 54/423, 13% [9.8-16%] | 20/265, 7.5% [4.8-12%] | 34/158, 22% [16-29%] | <0.001 | **0.003** |
| Fevers, including low-grade fevers | 80/423, 19% [15-23%] | 40/265, 15% [11-20%] | 40/158, 25% [19-33%] | 0.009 | 0.893 |
| Chills but no fever | 146/423, 35% [30-39%] | 69/265, 26% [21-32%] | 77/158, 49% [41-57%] | <0.001 | **<0.001** |
| Heat intolerance | 197/423, 47% [42-51%] | 101/265, 38% [32-44%] | 96/158, 61% [53-68%] | <0.001 | **<0.001** |
| Cold intolerance | 131/423, 31% [27-36%] | 68/265, 26% [21-31%] | 63/158, 40% [32-48%] | 0.002 | 0.211 |
| Night sweats | 175/423, 41% [37-46%] | 89/265, 34% [28-40%] | 86/158, 54% [46-62%] | <0.001 | **0.002** |

| | | | | | |
|---|---|---|---|---|---|
| Trouble falling or staying asleep | 303/423, 72% [67-76%] | 171/265, 65% [58-70%] | 132/158, 84% [77-89%] | <0.001 | **0.003** |
| Sleeping more than usual | 168/423, 40% [35-45%] | 96/265, 36% [30-42%] | 72/158, 46% [38-54%] | 0.057 | >0.999 |
| Nightmares | 73/423, 17% [14-21%] | 32/265, 12% [8.5-17%] | 41/158, 26% [19-34%] | <0.001 | **0.025** |
| Exercise intolerance | 333/423, 79% [74-82%] | 199/265, 75% [69-80%] | 134/158, 85% [78-90%] | 0.018 | >0.999 |
| Excessive fatigue | 369/423, 87% [84-90%] | 227/265, 86% [81-90%] | 142/158, 90% [84-94%] | 0.209 | >0.999 |
| Burning sensations | 116/423, 27% [23-32%] | 50/265, 19% [14-24%] | 66/158, 42% [34-50%] | <0.001 | **<0.001** |
| Tremors or shakiness | 161/423, 38% [33-43%] | 59/265, 22% [18-28%] | 102/158, 65% [57-72%] | <0.001 | **<0.001** |
| Tingling, pins and needles, numbness | 222/423, 52% [48-57%] | 109/265, 41% [35-47%] | 113/158, 72% [64-78%] | <0.001 | **<0.001** |
| Neuropathy (nerve sensations including pain) anywhere in the body | 192/423, 45% [41-50%] | 91/265, 34% [29-40%] | 101/158, 64% [56-71%] | <0.001 | **<0.001** |
| Seizures | 5/423, 1.2% [0.44-2.9%] | 3/265, 1.1% [0.29-3.5%] | 2/158, 1.3% [0.22-5.0%] | >0.999 | >0.999 |
| Abdominal pain | 136/423, 32% [28-37%] | 61/265, 23% [18-29%] | 75/158, 47% [40-56%] | <0.001 | **<0.001** |
| Acid reflux or heartburn | 123/423, 29% [25-34%] | 62/265, 23% [19-29%] | 61/158, 39% [31-47%] | <0.001 | 0.082 |
| Diarrhea | 132/423, 31% [27-36%] | 64/265, 24% [19-30%] | 68/158, 43% [35-51%] | <0.001 | **0.005** |
| Constipation | 108/423, 26% [21-30%] | 56/265, 21% [16-27%] | 52/158, 33% [26-41%] | 0.007 | 0.684 |

| | | | | | |
|---|---|---|---|---|---|
| Nausea/vomiting | 138/423, 33% [28-37%] | 72/265, 27% [22-33%] | 66/158, 42% [34-50%] | 0.002 | 0.185 |
| Loss of appetite | 129/423, 30% [26-35%] | 64/265, 24% [19-30%] | 65/158, 41% [33-49%] | <0.001 | **0.023** |
| Sore throat | 129/423, 30% [26-35%] | 76/265, 29% [23-35%] | 53/158, 34% [26-42%] | 0.293 | >0.999 |
| Congested or runny nose | 134/423, 32% [27-36%] | 76/265, 29% [23-35%] | 58/158, 37% [29-45%] | 0.086 | >0.999 |
| Palpitations (improper beating of the heart due to electrical impulse problems) | 222/423, 52% [48-57%] | 117/265, 44% [38-50%] | 105/158, 66% [58-74%] | <0.001 | **<0.001** |
| Bilateral neck throbbing around lymph nodes | 57/423, 13% [10-17%] | 20/265, 7.5% [4.8-12%] | 37/158, 23% [17-31%] | <0.001 | **<0.001** |
| Costochondritis (pain in the cartilage that connects a rib to the breastbone) | 105/423, 25% [21-29%] | 47/265, 18% [13-23%] | 58/158, 37% [29-45%] | <0.001 | **0.001** |
| Cough | 116/423, 27% [23-32%] | 73/265, 28% [22-33%] | 43/158, 27% [21-35%] | 0.941 | >0.999 |
| Coughing up blood | 7/423, 1.7% [0.73-3.5%] | 2/265, 0.8% [0.13-3.0%] | 5/158, 3.2% [1.2-7.6%] | 0.108 | >0.999 |
| Cold or burning feeling in lungs | 53/423, 13% [9.6-16%] | 26/265, 9.8% [6.6-14%] | 27/158, 17% [12-24%] | 0.029 | >0.999 |
| Difficulty swallowing | 74/423, 17% [14-22%] | 32/265, 12% [8.5-17%] | 42/158, 27% [20-34%] | <0.001 | **0.014** |
| Throat pain or discomfort | 95/423, 22% [19-27%] | 42/265, 16% [12-21%] | 53/158, 34% [26-42%] | <0.001 | **0.002** |
| Lump in throat | 58/423, 14% [11-17%] | 24/265, 9.1% [6.0-13%] | 34/158, 22% [16-29%] | <0.001 | **0.030** |
| Phlegm in back of throat | 102/423, 24% [20-29%] | 54/265, 20% [16-26%] | 48/158, 30% [23-38%] | 0.020 | >0.999 |

| | | | | | |
|---|---|---|---|---|---|
| Postnasal drip | 97/423, 23% [19-27%] | 56/265, 21% [16-27%] | 41/158, 26% [19-34%] | 0.254 | >0.999 |
| Runny nose | 74/423, 17% [14-22%] | 40/265, 15% [11-20%] | 34/158, 22% [16-29%] | 0.092 | >0.999 |
| Swollen lymph nodes | 82/423, 19% [16-24%] | 41/265, 15% [11-21%] | 41/158, 26% [19-34%] | 0.008 | 0.795 |
| Tachycardia (rapid heartbeat) at rest | 179/423, 42% [38-47%] | 89/265, 34% [28-40%] | 90/158, 57% [49-65%] | <0.001 | **<0.001** |
| Tachycardia (rapid heartbeat) after standing up | 194/423, 46% [41-51%] | 103/265, 39% [33-45%] | 91/158, 58% [49-65%] | <0.001 | **0.018** |
| Wheezing | 46/423, 11% [8.1-14%] | 21/265, 7.9% [5.1-12%] | 25/158, 16% [11-23%] | 0.012 | >0.999 |
| Shortness of breath or difficulty breathing | 246/423, 58% [53-63%] | 143/265, 54% [48-60%] | 103/158, 65% [57-72%] | 0.024 | >0.999 |
| Bone aches | 138/423, 33% [28-37%] | 65/265, 25% [20-30%] | 73/158, 46% [38-54%] | <0.001 | **<0.001** |
| Migraine | 129/423, 30% [26-35%] | 62/265, 23% [19-29%] | 67/158, 42% [35-51%] | <0.001 | **0.004** |
| Headache | 271/423, 64% [59-69%] | 149/265, 56% [50-62%] | 122/158, 77% [70-83%] | <0.001 | **0.001** |
| Calf cramps | 114/423, 27% [23-31%] | 54/265, 20% [16-26%] | 60/158, 38% [30-46%] | <0.001 | **0.008** |
| Pressure at base of head | 151/423, 36% [31-40%] | 71/265, 27% [22-33%] | 80/158, 51% [43-59%] | <0.001 | **<0.001** |
| Jaw pain | 92/423, 22% [18-26%] | 42/265, 16% [12-21%] | 50/158, 32% [25-40%] | <0.001 | **0.013** |
| Joint pain | 177/423, 42% [37-47%] | 88/265, 33% [28-39%] | 89/158, 56% [48-64%] | <0.001 | **<0.001** |
| Kidney pain | 35/423, 8.3% [5.9-11%] | 7/265, 2.6% [1.2-5.6%] | 28/158, 18% [12-25%] | <0.001 | **<0.001** |

| | | | | | |
|---|---|---|---|---|---|
| Mouth sores or sore tongue | 82/423, 19% [16-24%] | 38/265, 14% [10-19%] | 44/158, 28% [21-36%] | <0.001 | 0.064 |
| Muscle or body aches | 239/423, 57% [52-61%] | 129/265, 49% [43-55%] | 110/158, 70% [62-77%] | <0.001 | **0.003** |
| Persistent chest pain or pressure | 138/423, 33% [28-37%] | 68/265, 26% [21-31%] | 70/158, 44% [36-52%] | <0.001 | **0.007** |
| Painful scalp | 63/423, 15% [12-19%] | 23/265, 8.7% [5.7-13%] | 40/158, 25% [19-33%] | <0.001 | **<0.001** |
| Sharp or sudden chest pain | 131/423, 31% [27-36%] | 57/265, 22% [17-27%] | 74/158, 47% [39-55%] | <0.001 | **<0.001** |
| Changed sense of taste | 122/423, 29% [25-33%] | 66/265, 25% [20-31%] | 56/158, 35% [28-43%] | 0.021 | >0.999 |
| Changed sense of smell | 132/423, 31% [27-36%] | 66/265, 25% [20-31%] | 66/158, 42% [34-50%] | <0.001 | **0.028** |
| Floaters or flashes of light in vision | 117/423, 28% [24-32%] | 43/265, 16% [12-21%] | 74/158, 47% [39-55%] | <0.001 | **<0.001** |
| Loss of hearing | 37/423, 8.7% [6.3-12%] | 14/265, 5.3% [3.0-8.9%] | 23/158, 15% [9.6-21%] | 0.001 | 0.104 |
| Loss or decrease in quality of vision/blurry vision | 176/423, 42% [37-46%] | 90/265, 34% [28-40%] | 86/158, 54% [46-62%] | <0.001 | **0.003** |
| Phantom smells | 98/423, 23% [19-28%] | 43/265, 16% [12-21%] | 55/158, 35% [28-43%] | <0.001 | **0.001** |
| Phantom tastes | 36/423, 8.5% [6.1-12%] | 14/265, 5.3% [3.0-8.9%] | 22/158, 14% [9.1-21%] | 0.002 | 0.196 |
| Hallucinations (visual or auditory) | 26/423, 6.1% [4.1-9.0%] | 10/265, 3.8% [1.9-7.0%] | 16/158, 10% [6.1-16%] | 0.008 | 0.807 |
| Tinnitus or humming in ears | 193/423, 46% [41-51%] | 95/265, 36% [30-42%] | 98/158, 62% [54-70%] | <0.001 | **<0.001** |
| Skin bruising | 93/423, 22% [18-26%] | 42/265, 16% [12-21%] | 51/158, 32% [25-40%] | <0.001 | **0.008** |

| | | | | | |
|---|---|---|---|---|---|
| Change in nails (i.e. White spots, brittleness, change in moons) | 83/423, 20% [16-24%] | 35/265, 13% [9.5-18%] | 48/158, 30% [23-38%] | <0.001 | **0.002** |
| Tender or itchy rash or chilblains on the toes or foot) | 29/423, 6.9% [4.7-9.8%] | 5/265, 1.9% [0.70-4.6%] | 24/158, 15% [10-22%] | <0.001 | **<0.001** |
| Cracked or dry lips | 79/423, 19% [15-23%] | 35/265, 13% [9.5-18%] | 44/158, 28% [21-36%] | <0.001 | **0.018** |
| Dental problems (e.g., chipped tooth, tooth loss) | 54/423, 13% [9.8-16%] | 26/265, 9.8% [6.6-14%] | 28/158, 18% [12-25%] | 0.018 | >0.999 |
| Discoloration of the skin (for example: purple or blue on the hands or feet, no blistering) | 74/423, 17% [14-22%] | 28/265, 11% [7.3-15%] | 46/158, 29% [22-37%] | <0.001 | **<0.001** |
| Dry or peeling skin | 76/423, 18% [14-22%] | 35/265, 13% [9.5-18%] | 41/158, 26% [19-34%] | <0.001 | 0.091 |
| Dry scalp or dandruff | 61/423, 14% [11-18%] | 30/265, 11% [7.9-16%] | 31/158, 20% [14-27%] | 0.019 | >0.999 |
| Hair loss | 167/423, 39% [35-44%] | 79/265, 30% [24-36%] | 88/158, 56% [48-64%] | <0.001 | **<0.001** |
| Itchiness | 108/423, 26% [21-30%] | 46/265, 17% [13-23%] | 62/158, 39% [32-47%] | <0.001 | **<0.001** |
| Tender or itchy rash not on foot | 44/423, 10% [7.7-14%] | 21/265, 7.9% [5.1-12%] | 23/158, 15% [9.6-21%] | 0.031 | >0.999 |
| Chilblains (itching, bumps, red- to violet-colored patches on the hands or feet) | 24/423, 5.7% [3.7-8.4%] | 6/265, 2.3% [0.92-5.1%] | 18/158, 11% [7.1-18%] | <0.001 | **0.008** |
| Constant thirst | 111/423, 26% [22-31%] | 46/265, 17% [13-23%] | 65/158, 41% [33-49%] | <0.001 | **<0.001** |
| Changes in voice | 81/423, 19% [16-23%] | 46/265, 17% [13-23%] | 35/158, 22% [16-30%] | 0.226 | >0.999 |
| Clogged ears | 98/423, 23% [19-28%] | 49/265, 18% [14-24%] | 49/158, 31% [24-39%] | 0.003 | 0.299 |

| | | | | | |
|---|---|---|---|---|---|
| Dizziness | 249/423, 59% [54-64%] | 131/265, 49% [43-56%] | 118/158, 75% [67-81%] | <0.001 | **<0.001** |
| Dry eyes | 120/423, 28% [24-33%] | 56/265, 21% [16-27%] | 64/158, 41% [33-49%] | <0.001 | **0.002** |
| Fatigue | 361/423, 85% [82-89%] | 219/265, 83% [77-87%] | 142/158, 90% [84-94%] | 0.042 | >0.999 |
| Irregular or skipped menstrual cycles | 78/423, 18% [15-23%] | 38/265, 14% [10-19%] | 40/158, 25% [19-33%] | 0.005 | 0.462 |
| Menstrual cycles that are heavier or lighter than normal | 81/423, 19% [16-23%] | 40/265, 15% [11-20%] | 41/158, 26% [19-34%] | 0.006 | 0.575 |
| New allergies | 68/423, 16% [13-20%] | 32/265, 12% [8.5-17%] | 36/158, 23% [17-30%] | 0.004 | 0.354 |
| Inability to eat or tolerate food | 90/423, 21% [18-26%] | 39/265, 15% [11-20%] | 51/158, 32% [25-40%] | <0.001 | **0.002** |
| Swollen hands or feet | 78/423, 18% [15-23%] | 36/265, 14% [9.8-18%] | 42/158, 27% [20-34%] | <0.001 | 0.081 |
| Fainting | 43/423, 10% [7.5-14%] | 13/265, 4.9% [2.7-8.4%] | 30/158, 19% [13-26%] | <0.001 | **<0.001** |
| Weakened neck | 69/423, 16% [13-20%] | 23/265, 8.7% [5.7-13%] | 46/158, 29% [22-37%] | <0.001 | **<0.001** |
| [1] Pearson's Chi-squared test; Fisher's exact test | | | | | |
| [2] Bonferroni correction for multiple testing | | | | | |
| CI, confidence interval | | | | | |

**eTable 5. Variable importance in the machine learning models when including all symptom variables**

| Symptom | GBM Percent Importance | XGB Mean Absolute Shapley Value | XGB Gain Percent Importance |
|---|---|---|---|
| Tremors or shakiness | 27.45 | 0.510490907 | 44.85 |
| Floaters or flashes of light in vision | 8.44 | 0.139807497 | 10.16 |
| Hair loss | 5.27 | 0.121706726 | 5.26 |
| Tingling, pins and needles, numbness | 4.47 | 0.166079321 | 9.82 |
| Tinnitus or humming in ears | 3.75 | 0.120033087 | 4.98 |
| Sharp or sudden chest pain | 3.46 | 0.073050261 | 4.94 |
| Abdominal pain | 2.98 | 0.018376863 | 0.97 |
| Itchiness | 2.82 | 0.013954319 | 0.78 |
| Migraine | 2.46 | 0 | 0 |
| Kidney pain | 2.23 | 0.04543798 | 4.6 |
| Sore throat | 2.09 | 0.014627527 | 0.68 |
| Painful scalp | 1.98 | 0 | 0 |
| Dizziness | 1.94 | 0.056787718 | 2.29 |
| Cough | 1.79 | 0 | 0 |
| Tender or itchy rash on toes or foot | 1.76 | 0.008380492 | 1.06 |
| Acid reflux or heartburn | 1.72 | 0 | 0 |
| Postnasal drip | 1.68 | 0 | 0 |
| Constant thirst | 1.65 | 0.01901079 | 0.73 |
| Tachycardia at rest | 1.6 | 0.021019243 | 0.98 |
| Headache | 1.45 | 0.018371266 | 0.7 |
| Phantom smells | 1.34 | 0 | 0 |
| Inability to eat or tolerate food | 1.27 | 0 | 0 |
| Changes in voice | 1.2 | 0 | 0 |
| Constipation | 1.18 | 0 | 0 |
| Costochondritis | 1.17 | 0 | 0 |
| Calf cramps | 1.06 | 0.016288212 | 1.09 |
| Trouble falling/staying asleep | 1.01 | 0.022355786 | 0.72 |
| Neuropathy | 0.95 | 0.038334172 | 1.46 |
| Wheezing | 0.85 | 0 | 0 |
| Bilateral neck throbbing | 0.81 | 0 | 0 |
| Difficulty swallowing | 0.68 | 0 | 0 |
| Loss of hearing | 0.66 | 0 | 0 |

| | | | |
|---|---|---|---|
| Tachycardia after standing up | 0.64 | 0 | 0 |
| Mouth sores or sore tongue | 0.6 | 0 | 0 |
| Feelings of impending doom | 0.59 | 0 | 0 |
| Phlegm in back of throat | 0.59 | 0 | 0 |
| Throat pain | 0.55 | 0 | 0 |
| Cold intolerance | 0.51 | 0 | 0 |
| Clogged ears | 0.51 | 0 | 0 |
| Chilblains on hands or feet | 0.5 | 0.009202775 | 0.86 |
| Fainting | 0.49 | 0 | 0 |
| Excessive fatigue | 0.43 | 0 | 0 |
| Heat intolerance | 0.41 | 0 | 0 |
| Chills | 0.34 | 0 | 0 |
| Difficulty speaking properly | 0.33 | 0 | 0 |
| Diarrhea | 0.31 | 0 | 0 |
| Anxiety | 0 | 0 | 0 |
| Confusion | 0 | 0 | 0 |
| Brain fog | 0 | 0 | 0 |
| Memory problems | 0 | 0 | 0 |
| Suicidal thoughts | 0 | 0 | 0 |
| Abnormally low temperature | 0 | 0 | 0 |
| Fevers | 0 | 0 | 0 |
| Night sweats | 0 | 0 | 0 |
| Sleeping more than usual | 0 | 0 | 0 |
| Nightmares | 0 | 0 | 0 |
| Exercise intolerance | 0 | 0 | 0 |
| Burning sensations | 0 | 0 | 0 |
| Seizures | 0 | 0 | 0 |
| Nausea/vomiting | 0 | 0 | 0 |
| Loss of appetite | 0 | 0 | 0 |
| Congested or runny nose | 0 | 0 | 0 |
| Palpitations | 0 | 0 | 0 |
| Coughing up blood | 0 | 0 | 0 |
| Cold or burning feeling in lungs | 0 | 0 | 0 |
| Lump in throat | 0 | 0 | 0 |
| Runny nose | 0 | 0 | 0 |
| Swollen lymph nodes | 0 | 0 | 0 |
| Shortness of breath | 0 | 0 | 0 |
| Bone aches | 0 | 0 | 0 |

| | | | |
|---|---|---|---|
| Pressure at base of head | 0 | 0 | 0 |
| Jaw pain | 0 | 0 | 0 |
| Joint pain | 0 | 0 | 0 |
| Muscle or body aches | 0 | 0.014623417 | 0.72 |
| Persistent chest pain or pressure | 0 | 0 | 0 |
| Changed sense of taste | 0 | 0 | 0 |
| Changed sense of smell | 0 | 0.014390654 | 0.69 |
| Loss or decrease in quality of vision/blurry vision | 0 | 0 | 0 |
| Phantom tastes | 0 | 0 | 0 |
| Hallucinations | 0 | 0 | 0 |
| Skin bruising | 0 | 0 | 0 |
| Change in nails | 0 | 0.013030063 | 0.69 |
| Cracked or dry lips | 0 | 0 | 0 |
| Dental problems | 0 | 0 | 0 |
| Discoloration of skin | 0 | 0 | 0 |
| Dry skin | 0 | 0 | 0 |
| Dry scalp | 0 | 0 | 0 |
| Tender or itchy rash not on foot | 0 | 0 | 0 |
| Dry eyes | 0 | 0 | 0 |
| Fatigue | 0 | 0 | 0 |
| New allergies | 0 | 0 | 0 |
| Swollen hands/feet | 0 | 0 | 0 |
| Weakened neck | 0 | 0.01949914 | 0.97 |
| Abbreviations: GBM, gradient-boosting machine using a permutation-based approach; XGB Gain, XGBoost tree machine learning model with gain in accuracy metric. | | | |
| In this table, symptoms were sorted based on their importance in the GBM method. For each of the three methods used, we sorted the variables based on their importance and, using this fixed sorting, progressively excluded those with least importance from the model by evaluating the change in the AUC. | | | |

**eMethods 1. Health status and symptom severity questions**

1. Please choose one point in this 0-100 scale, which can best represent your health today (0 means the worst and 100 means the best). [“slider”]
2. We are trying to get a sense of how bad your long COVID symptoms are when you feel them the most. On the slider below, with 0 being a trivial illness and 100 being unbearable, please let us know what the worst days are like. ["slider"]

**eMethods 2. Pre-pandemic comorbidities questions**

Have you ever been told by a doctor before January 2020 that you have any of the following?
Check all that apply
["multiple choice"]

1. Any allergies
2. Arthritis (including rheumatoid arthritis, gout, lupus, or fibromyalgia)
3. Asthma
4. Autoimmune disease (including lupus, scleroderma, etc.)
5. Bleeding disorder (including sickle cell disease or Thalassemia)
6. Blood clots
7. Cancer or malignancy of any kind
8. Cerebrovascular conditions affecting blood vessels to or in the brain (including stroke)
9. Chronic lung disease (including emphysema, chronic bronchitis, chronic obstructive pulmonary disease (COPD), or pulmonary fibrosis)
10. Cystic fibrosis
11. Diabetes
12. Ehlers Danlos Syndrome (hypermobile joints)
13. Gastrointestinal issues (including IBS or acid reflux)
14. Heart attack, also called myocardial infarction
15. Heart conditions (including coronary artery disease or cardiomyopathies)
16. Heart failure
17. High cholesterol
18. History of organ transplant (including kidney, liver, heart, or lung)
19. Hypertension or high blood pressure
20. Immunocompromised state (including weakened immune system from blood or bone marrow transplant, immune deficiencies, HIV, use of corticosteroids, or use of other immune-weakening medicines)
21. Kidney disease
22. Liver disease
23. Lyme disease
24. MCAS (mast cell activation syndrome) or other mast cell disorders
25. ME/CFS (myalgic encephalomyelitis/chronic fatigue syndrome)
26. Migraines
27. Neurologic conditions (including seizures, dementia, multiple sclerosis, Parkinson's, neuropathy, small fiber neuropathy, etc.)
28. Postural orthostatic hypotension (POTS) or dysautonomia
29. Spinal disorder(s)
30. Tremors/Internal vibrations
31. Other
32. None of the above

Have you ever been told by a doctor before January 2020 that you have any of the following?
Check all that apply
["multiple choice"]

1. Depressive disorders
2. Anxiety disorders
3. Schizophrenia spectrum and other psychotic disorders
4. Bipolar and related disorders
5. Obsessive-compulsive and related disorders
6. Trauma- and stressor-related disorders
7. Feeding and eating disorders
8. Somatic symptoms (excessive thoughts, feelings and behaviors relating to the physical symptoms) and related disorders
9. Other
10. None of the above

**eMethods 3. Current conditions questions**

Currently, have you ever been told by a doctor that you have any of the following? Check all that apply ["multiple choice"]

1. Any allergies
2. Arthritis (including rheumatoid arthritis, gout, lupus, or fibromyalgia)
3. Asthma
4. Autoimmune disease (including lupus, scleroderma, etc.)
5. Bleeding disorder (including sickle cell disease or Thalassemia)
6. Blood clots
7. Cancer or malignancy of any kind
8. Cerebrovascular conditions affecting blood vessels to or in the brain (including stroke)
9. Chronic lung disease (including emphysema, chronic bronchitis, chronic obstructive pulmonary disease (COPD), or pulmonary fibrosis)
10. Cystic fibrosis
11. Diabetes
12. Ehlers Danlos Syndrome (hypermobile joints)
13. Gastrointestinal issues (including IBS or acid reflux)
14. Heart attack, also called myocardial infarction
15. Heart conditions (including coronary artery disease or cardiomyopathies)
16. Heart failure
17. High cholesterol
18. History of organ transplant (including kidney, liver, heart, or lung)
19. Hypertension or high blood pressure
20. Immunocompromised state (including weakened immune system from blood or bone marrow transplant, immune deficiencies, HIV, use of corticosteroids, or use of other immune-weakening medicines)
21. Kidney disease
22. Liver disease
23. Long COVID
24. Lyme disease
25. MCAS (mast cell activation syndrome) or other mast cell disorders
26. ME/CFS (myalgic encephalomyelitis/chronic fatigue syndrome)
27. Migraines
28. Neurological conditions (including seizures, dementia, multiple sclerosis, Parkinson's, neuropathy, small fiber neuropathy, etc.)
29. Postural orthostatic hypotension (POTS) or dysautonomia
30. Spinal disorder(s)
31. Vaccine injury
32. Other
33. None of the above

Currently, have you ever been told by a doctor that you have any of the following? Check all that apply ["multiple choice"]

1. Depressive disorders
2. Anxiety disorders
3. Schizophrenia spectrum and other psychotic disorders
4. Bipolar and related disorders
5. Obsessive-compulsive and related disorders
6. Trauma- and stressor-related disorders
7. Feeding and eating disorders
8. Somatic symptom and related disorders
9. Other
10. None of the above

**eMethods 4. Long COVID symptoms questions**

Please select all following health conditions that you have had as a result of long COVID. Check all that apply ["multiple choice"]

1. Abnormally low temperature
2. Fevers, including low-grade fevers
3. Chills but no fever
4. Heat intolerance
5. Cold intolerance
6. Night sweats
7. Other
8. None of the above

Please select all following health conditions that you have had as a result of long COVID. Check all that apply ["multiple choice"]

1. Trouble falling or staying asleep
2. Sleeping more than usual
3. Nightmares
4. Exercise intolerance
5. Excessive fatigue
6. Other
7. None of the above

Please select all following health conditions that you have had as a result of long COVID. Check all that apply ["multiple choice"]

1. Burning sensations
2. Tremors or shakiness
3. Internal tremors or buzzing/vibration
4. Tingling, pins and needles, numbness
5. Neuropathy (nerve sensations including pain) anywhere in the body
6. Seizures
7. Other
8. None of the above

Please select all following health conditions that you have had as a result of long COVID. Check all that apply ["multiple choice"]

1. Abdominal pain
2. Acid reflux or heartburn
3. Diarrhea
4. Constipation
5. Nausea/Vomiting
6. Loss of appetite
7. Other
8. None of the above

Please select all following health conditions that you have had as a result of long COVID. Check all that apply ["multiple choice"]

1. Sore throat
2. Congested or runny nose
3. Palpitations (improper beating of the heart due to electrical impulse problems)
4. Bilateral neck throbbing around lymph nodes

5. Costochondritis (pain in the cartilage that connects a rib to the breastbone)
6. Cough
7. Coughing up blood
8. Cold or burning feeling in lungs
9. Difficulty swallowing
10. Throat pain or discomfort
11. Lump in throat
12. Phlegm in back of throat
13. Postnasal drip
14. Runny nose
15. Swollen lymph nodes
16. Tachycardia (rapid heartbeat) at rest
17. Tachycardia (rapid heartbeat) after standing up
18. Wheezing
19. Shortness of breath or difficulty breathing
20. Other
21. None of the above

Please select all following health conditions that you have had as a result of long COVID. Check all that apply ["multiple choice"]

1. Bone aches
2. Migraine
3. Headache
4. Calf cramps
5. Pressure at base of head
6. Jaw pain
7. Joint pain
8. Kidney pain
9. Mouth sores or sore tongue
10. Muscle or body aches
11. Persistent chest pain or pressure
12. Painful scalp
13. Sharp or sudden chest pain
14. Other
15. None of the above

Please select all following health conditions that you have had as a result of long COVID. Check all that apply ["multiple choice"]

1. Changed sense of taste
2. Changed sense of smell
3. Floaters or flashes of light in vision
4. Loss of hearing
5. Loss or decrease in quality of vision/blurry vision
6. Phantom smells
7. Phantom tastes
8. Hallucinations (visual or auditory)
9. Tinnitus or humming in ears
10. Other

11. None of the above

Please select all following health conditions that you have had as a result of long COVID. Check all that apply ["multiple choice"]

1. Skin bruising
2. Change in nails (i.e. white spots, brittleness, change in moons)
3. Tender or itchy rash or chilblains on the toes or foot)
4. Cracked or dry lips
5. Dental problems (e.g., chipped tooth, tooth loss)
6. Discoloration of the skin (for example: purple or blue on the hands or feet, no blistering)
7. Dry or peeling skin
8. Dry scalp or dandruff
9. Hair loss
10. Itchiness
11. Tender or itchy rash not on foot
12. Chilblains (itching, bumps, red- to violet-colored patches on the hands or feet)
13. Other
14. None of the above

Please select all following health conditions that you have had as a result of long COVID. Check all that apply ["multiple choice"]

1. Constant thirst
2. Changes in voice
3. Clogged ears
4. Dizziness
5. Dry eyes
6. Fatigue
7. Irregular or skipped menstrual cycles
8. Menstrual cycles that are heavier or lighter than normal
9. New allergies
10. Inability to eat or tolerate food
11. Swollen hands or feet
12. Fainting
13. Weakened neck
14. Other
15. None of the above

Please select all that you have. Check all that apply ["multiple choice"]

1. Anxiety
2. Confusion
3. Brain fog; difficulty concentrating or focusing
4. Feelings of impending doom
5. Memory problems
6. Difficulty speaking properly
7. Suicidal thoughts
8. Other
9. None of the above

**eMethods 5. RECOVER and LISTEN questions**

| Symptom | Score | Symptom as it appears in the RECOVER questionnaire and severity criteria, if applicable[a] | Symptom as it appears in the LISTEN questionnaire |
|---|---|---|---|
| Smell/taste | 8 | • Loss of or change in smell or taste | • Changed sense of taste<br>• Changed sense of smell<br>• Phantom smells<br>• Phantom tastes |
| Postexertional malaise | 7 | • Post-exertional malaise (Symptoms worse after even minor physical or mental effort) | • Exercise intolerance |
| Chronic cough | 4 | • Persistent (chronic) cough | • Cough |
| Brain fog[c] | 3 | • Problems thinking or concentrating ("brain fog")[c]<br>o Neuro-QoL Cognition Score (≤40) | • Brain fog; difficulty concentrating or focusing[c] |
| Thirst | 3 | • Excessive thirst | • Constant thirst |
| Palpitations | 2 | • Palpitations, racing heart, arrhythmia, skipped beats | • Palpitations (improper beating of the heart due to electrical impulse problems) |
| Chest pain[c] | 2 | • Chest pain (including chest tightness, pressure)[c]<br>o SAQ-7 (<75) | • Persistent chest pain or pressure[c] |
| Fatigue[c] | 1 | • Fatigue (being very tired)[c]<br>o PROMIS Fatigue Score (moderate or worse) | • Excessive fatigue[c] |
| Sexual desire or capacity | 1 | • Changes in desire for, comfort with or capacity for sex | Not asked in LISTEN |
| Dizziness | 1 | • Feeling faint, dizzy, "goofy"; difficulty thinking soon after standing up from a sitting or lying position | • Dizziness |

| | | | |
|---|---|---|---|
| Gastrointestinal | 1 | • Gastrointestinal (belly) symptoms (feeling full or vomiting after eating, diarrhea, constipation) | • Inability to eat or tolerate food<br>• Nausea/Vomiting<br>• Diarrhea<br>• Constipation |
| Abnormal movements | 1 | • Abnormal movements | Not asked in LISTEN[d] |
| Hair loss | 0 | • Hair loss | • Hair loss |

Abbreviations: LISTEN= Listen to Immune, Symptom, and Treatment Experiences Now, Neuro-QoL= Quality of Life in Neurological Disorders, PROMIS=Patient-Reported Outcomes Measurement Information System, RECOVER=Researching COVID to Enhance Recovery, SAQ=Seattle Angina Questionnaire.

a) The table was adapted from Table 2 and eTable 1 of the paper Thaweethai T, Jolley SE, Karlson EW, et al. Development of a Definition of Postacute Sequelae of SARS-CoV-2 Infection. *JAMA.* 2023;329(22):1934–1946. doi:10.1001/jama.2023.8823

b) For RECOVER symptoms that corresponded to multiple LISTEN symptoms, a LISTEN participant receives the corresponding score if they reported at least one of the symptom choices. For example, a participant received a score of 8 if they reported one or more of the four symptoms of 1) Changed sense of taste, 2) Changed sense of smell, 3) Phantom smells, or 4) Phantom tastes

c) For RECOVER symptoms that require additional severity criteria, LISTEN did not assess severity using the same instruments as RECOVER.

d) RECOVER assessed abnormal movements, tremor, seizures, and paralysis as separate symptoms. LISTEN assessed tremors/shakiness and internal tremors or buzzing/vibration but did not assess abnormal movements.
